# Unequal impact of the Covid-19 pandemic on excess deaths, life expectancy, and premature mortality across Spanish regions in 2020 and 2021

**DOI:** 10.1101/2021.11.29.21266617

**Authors:** Nazrul Islam, Fernando J. García López, Dmitri A. Jdanov, Miguel Ángel Royo- Bordonada, Kamlesh Khunti, Sarah Lewington, Ben Lacey, Martin White, Eva JA Morris, María Victoria Zunzunegui

## Abstract

Spain is one of the most heavily affected countries by the Covid-19 pandemic. In this study, we estimated the regional inequalities in excess deaths and premature mortality in Spain. Between January 2020 and June 2021, an estimated 89,200 (men: 48,000; women: 41,200) excess deaths occurred in the 17 Spanish regions with a substantial variability (highest in Madrid: 22,000, lowest in Canary Islands: -210). Highest reductions in life expectancy at birth (*e*_0_) in 2020 were observed in Madrid (men: -3.48 years, women: -2.15), Castile La Mancha (men: -2.67, women: -2.30), and Castile and León (men: -2.00, women: -1.32). In the first six months of 2021, the highest reduction in *e*_0_ was observed in Valencian Community (men: -2.04, women: -1.63), Madrid (men: - 2.37), and Andalusia (men: -1.75; women: -1.43). In some Spanish regions, life expectancy at age 65 during the Covid-19 pandemic in 2020 was comparable to that observed as far back as 20 years ago.

## Introduction

Since the emergence of Covid-19, countries and jurisdictions have employed a wide range of public health policy interventions with a view to minimising the impact of the pandemic.[1–7] These measures have affected many socioeconomic determinants of health,[8–11] including the provision of healthcare services.[12–15] Measuring the overall impact of the Covid-19 pandemic presents a multitude of challenges, including differences in the accuracy and completeness of reported deaths from Covid-19.[2,16] Therefore, ‘excess deaths’ (observed minus expected deaths) is widely considered the gold standard in estimating the overall impact of the pandemic since robust data on all-cause mortality is less sensitive to misclassification in designating the cause of deaths.[15,17–21] We have previously reported large differences between reported Covid-19 deaths and estimated excess deaths associated with the Covid-19 pandemic in 2020 at a national level,[22] but variations in excess deaths within countries at a regional level has not been well described.

However, ‘excess deaths’ does have limitations as a measure of impact. In particular, it does not account for the age at death.[23–25] Analysis of life expectancy (LE) and Years of Life Lost (YLL) provides a more granular estimation of premature mortality. Life expectancy is an indication of the average number of years that people can expect to survive if the age-specific death rates of that year remain unchanged for the remainder of their life.[26–28] YLL takes into account the age distributions at death by giving larger weights to deaths at younger ages.[24] While LE is a widely used standardised measure based on a synthetic life table cohort, YLL is based on the numbers of deaths observed in real populations. An analysis of excess deaths, LE, and YLL provides a comprehensive examination on the effects of the Covid-19 pandemic.[22,25] For example, we previously showed that, with a very similar excess death rates (per 100,000) in the US (160) and Spain (161),[22] excess YLL (per 100□000) was substantially higher in the US (3400) compared to Spain (1900).[25] This analysis revealed that the US had a markedly increased mortality rates in people <65 years in 2020 than Spain.[25]

Spain, a country composed of 17 autonomous regions which vary markedly in economic and social conditions, has been heavily affected by Covid-19.[29] Each region has its own department of health with responsibility for governance and organization of health and social services. Overall coordination of the 17 departments of health and the national monitoring of health system performance is assured at the national level by the central government of the State, guided by the core principles of universality, free access, equity, and financial fairness on which the Spanish health system is based. Financing is mostly based on general taxation, but there is wide variability in public health expenditures per capita across regions.[30] There is evidence of some heterogeneity in the accuracy and completeness of reported deaths from Covid-19 across regions, particularly during the first months,[27] when some regional health systems almost collapsed.[31] Measures taken to respond to the Covid-19 pandemic have also varied substantially across regions.[32] An analysis of regional differences in excess deaths, LE, and YLL could identify important inequities in the impact of the Covid-19 pandemic and thus informing future public health practices and pandemic preparedness.

We aimed to report excess deaths, changes in life expectancy, and years of life lost from the Covid-19 pandemic in the regions of Spain, accounting for temporal trends and seasonal variations in mortality within regions during January 2020 through June 2021. This work applies previously developed methodology to estimate the impact on mortality of the Covid-19 pandemic on excess deaths, LE, and YLL.[22,25]

## Methods

### Study design & eligibility

This is a time-series analysis using the annual all-cause mortality on the whole population data obtained from Spain between 2010 and 2021, disaggregated by autonomous Spanish regions, age, and sex. We excluded two small Spanish territories, Ceuta and Melilla, since they are African cities with small populations which would result in unstable estimates.

### Source of data

Our analysis required weekly data, disaggregated by Spanish regions, age, and sex, for the estimation of excess deaths.[22] We obtained weekly death data by 5-year age groups up to 90 years of age or older, sex and region from the Spanish National Statistics Office. Weekly mortality data from first week of 2019 and to week 24 of 2021 are publicly available at *https://www.ine.es/jaxiT3/Tabla.htm?t=35179*. Previous research showed that comparing the pandemic estimates with a single year in the pre-pandemic period ignores the recent trend in health improvements and seasonal variability.[25] We therefore obtained additional granular weekly data by region, age and sex for 2015-2018 from the National Statistics Offices upon request.

We also obtained monthly mortality data by five-year age groups (including infant mortality), sex and region between January 2010 and December 2020, which enabled us to use a longer reference period for the calculation of changes in life expectancy and years of life lost.

We obtained the annual population figures by age, sex, and region from the continuous census of the National Statistics Office (*https://www.ine.es/jaxiT3/Tabla.htm?t=10262&L=1*). All annual populations were 1 July estimates, except for 2021, when 1 January estimates were used.

### Statistical analysis

#### Estimation of excess deaths

We used our previously developed validated methodology for the estimation of excess deaths, extensive details of which have been published elsewhere.[22,33] In summary, observed weekly deaths in 2020 in each strata (by age, sex, and regions) were compared to the stratum-specific number of expected deaths. Excess death was the difference between the observed and the expected deaths. Expected death has been estimated based on the historical trends (2015-2019) using an over-dispersed Poisson model that accounts for temporal trends, seasonal and natural variability in mortality.[22,33]

Specifically, our mean model is:

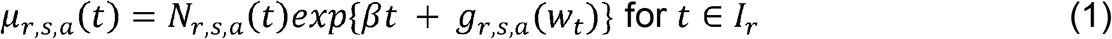

Here, *μ*_*r,s,a*_ (*t*) is the expected number of deaths at region *r*, sex *s*, age group *a* at week *t. N*_*r,s,a*_(*t*) is an offset term for the population size, *β* represents an estimated linear effect of time to account for slow-moving changes in mortality, *g*_*r,s,a*_(*w*_*t*_) is a function to account for seasonal trends, where *w*_*t*_ ∈ {1,…,52} represents the weeks of the year, and *I*_*r*_ is a region-specific reference interval (here, 2015-2019 for all regions) used to fit the model.

We calculated the smooth estimates of weekly percent change using:

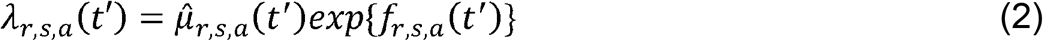

In equation (2), *λ*_*r,s,a*_(*t′*) and 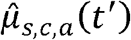 represent the average number of observed and counterfactual deaths at the respective stratum in week *t′*, respectively, and *f*_*r,s,a*_(*t′*) is a natural cubic spline with 3 internal knots per calendar year (which returned the lowest median of the median absolute deviation in the reference period). The smooth estimate of weekly percent change from average at 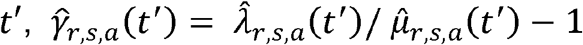. We used excessmort R package to fit our modelling scheme, and additional details, including the smooth estimates of the weekly percent change and the estimation of the uncertainty around the point estimates, have been described extensively elsewhere.[22,33]

To avoid a false sense of precision,[21,22] numbers <1000, between 1000 and <100000, and those ≥100000 were rounded to the nearest ten, hundred, and thousand, respectively. Excess deaths were directly standardised using the 2013 European Standard Population.[34]

The estimated number of excess deaths were compared with the reported number of confirmed Covid-19 deaths in each region reported by the autonomous region to the National Epidemiology Center as of June 30, 2021.

### Calculation of life expectancy and YLL in 2020 and 2021

The standard algorithm for calculating (abridged) life tables assumes that data are available for the first year of life and for the first five years of life. We have obtained annual data by granular age (including infant mortality) and sex groups for each region for 2010-2020. The data we have obtained for 2021 had similar granularity except that the death counts were available for 0-4 age groups without further disaggregation into <1, and 1-4 as required for the calculation of life expectancy. Using a similar approach, we have applied the following method to split the first age group. First, we split population exposures using proportions from previous years:

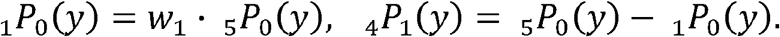

Here _*a*_*P*_*b*_(*y*) denotes population exposures in year *y* in age interval [*b*;*b* + *a*) and is *w*_1_ is average proportion of age group <1:

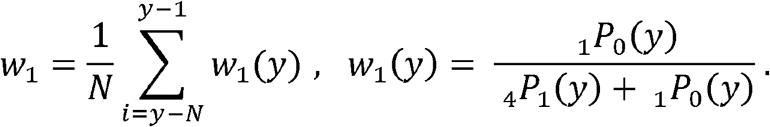

At the next step, we estimate infant mortality rate (IMR) in year *y* using linear regression fitted using IMRs from *N* previous years. The number of deaths in the first year of Life _1_*D*_0_ is calculated as follows:

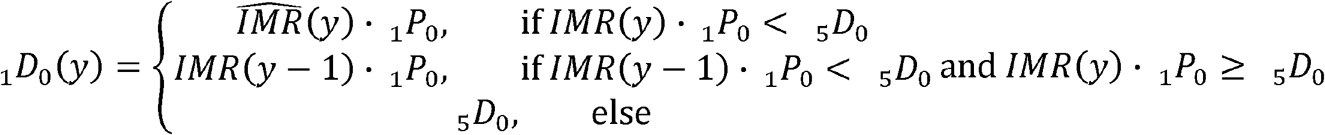

And the number of deaths in age interval [1,5) is

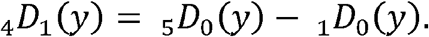

We applied this algorithm separately to each sex in each region.

We have validated this approach extensively,[25] and found that the difference was <0.01 years for all ages below 80 (maximum absolute difference was 0.02). In this analysis, we have validated our approach for 2020 data, and found that the mean difference was 0.10 years (median: 0.10, maximum: 0.19).

We used standard life table methodology to estimate life expectancy at birth and at age 65.[35,36] The details of the methodology have been published elsewhere.[25] In summary, we converted the death rates, _*n*_*m*_*x*_, into probabilities of death, _*n*_*q*_*x*_, where index *n* refers to the length of the interval, and *x* denotes the beginning of the age interval. For example, _*n*_*q*_*x*_ denotes the probability of death in age interval [*x, x + n*).

Let, _*n*_*q*_*x*_ be the average number of years lived within the age interval [*x, x + n*) for people dying at that age. We assume that 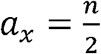 for all single-year ages except age 0 (see below). We then compute _*n*_*q*_*x*_ from _*n*_*m*_*x*_ and _*n*_*a*_*x*_ according to the formula,

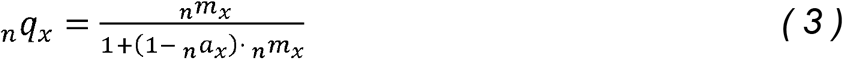

for *x* = 0,1,5,10,…,85. For the open age interval (90+), we set 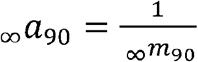 and _∞_*q*_90_ = 1

For infants <1 year of age, we used the formulas for *a*_0_ suggested by Preston *et al*.,[35] which are adapted from the Coale-Demeny model life tables.[36] Thus, if *m*_0_ ≥ 0.107:

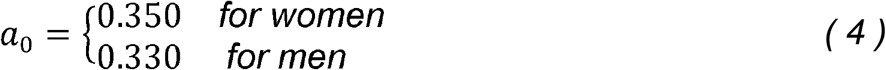

On the other hand, if *m*_0_ ≥ 0.107:

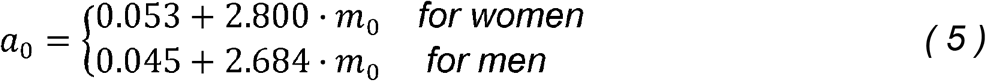

To complete the life table calculation, let _*n*_*p*_*x*_ be the probability of surviving from age *x* to *x* + *n*.Therefore,

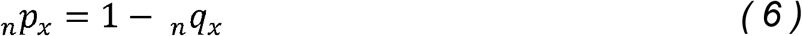

for all ages *x*. Let the radix (the starting number of new-borns) of the life table be *l*_0_ = 100,000. Then, the number of survivors (out of 100,000) at age *x* is

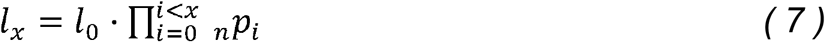

The distribution of deaths by age in the life-table population is

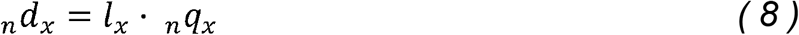

for *x =* 0,1,5,…,85. For the open age category, _∞_*d*_x90 =_ *l*_90._

The person-years lived by the life-table population in the age interval [*x, x + n*) are

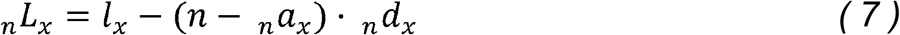

for *x =* 0,1,5,…,85. For the open age category,_∞_*L*_90 =_. *l*_90_.*a*_*90*_ The person-years remaining for individuals of age *x* equal

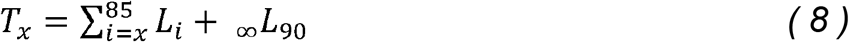

for *x =* 0,1,5,…,85. Remaining life expectancy at age *x* is

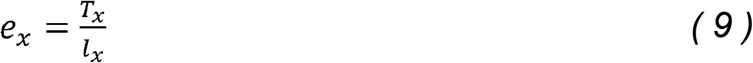

for *x =* 0,1,5,…,85, *e*_90_ = 1/_∞_*m*_90_

To attribute an equal loss of lifetime produced by deaths at the same age across the regions,[37,38] we calculated the YLL using World Health Organization (WHO) standard life table, as is used in the Global Burden of Disease, Injuries and Risk Factor (GBD) study.[38,39] We estimated the YLL with the following equation:

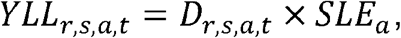

where *D*_*r,s,a,t*_ is the number of deaths in region *r*, sex *s*, age *a*, and calendar year *t*, and *SLE*_*a*_ is the WHO Global Health Estimates standard life expectancy at age *a*, [38]

### Calculation of changes in life expectancy and YLL in 2020

Within each region, sex, and age groups, the change in life expectancy at birth was estimated as the difference between the observed and ‘expected’ life expectancy at birth (*e*_0_) and age 65 (*e*_65_) in 2020. The expected life expectancy for 2020 was estimated based on Lee-Carter forecasting using 2010-2019 data.[40] Similar methodology was used to calculate the changes in YLL in 2020 within each region, sex, and age groups.

We applied bootstrap methods to estimate the statistical uncertainty from the sample of 5000 iterations. Once a distribution of age-and-sex-specific forecasted mortality rates has been derived, we generated a random set of death rates and calculated our parameters of interest (LE, YLL, and changes in LE and YLL). The 2.5^th^ and the 97.5^th^ quantile of the bootstrap distribution were used as the 95% confidence intervals for each region and sex strata.

We have used the same methodology to also estimate the changes in LE and YLL for 2021 using data up to the 24^th^ week of 2021. Statistical analyses were conducted using R (version 4.1.0). The Lee Carter forecast was performed using the R-package *demography*.[54] Ethics approval was waived by the COMITÉ DE ÉTICA DE LA INVESTIGACIÓN (No: CEI PI 77_2021) of Instituto de Salud Carlos III because all data analysed in this study were fully anonymised and aggregated without any identifiable information.

## Results

### Excess deaths during 2020-2021

Relative to the trend observed between 2000 and 2019, the age-specific and age-standardised death rates increased sharply in 2020, with a steeper increase in the elderly. The increase was particularly higher in Madrid, Castile La Mancha, Castile and León, and Catalonia with a greater increase in men than women (Figure 1(A-D) and Supplementary Figure 1(A-H).

**Figure 1.**
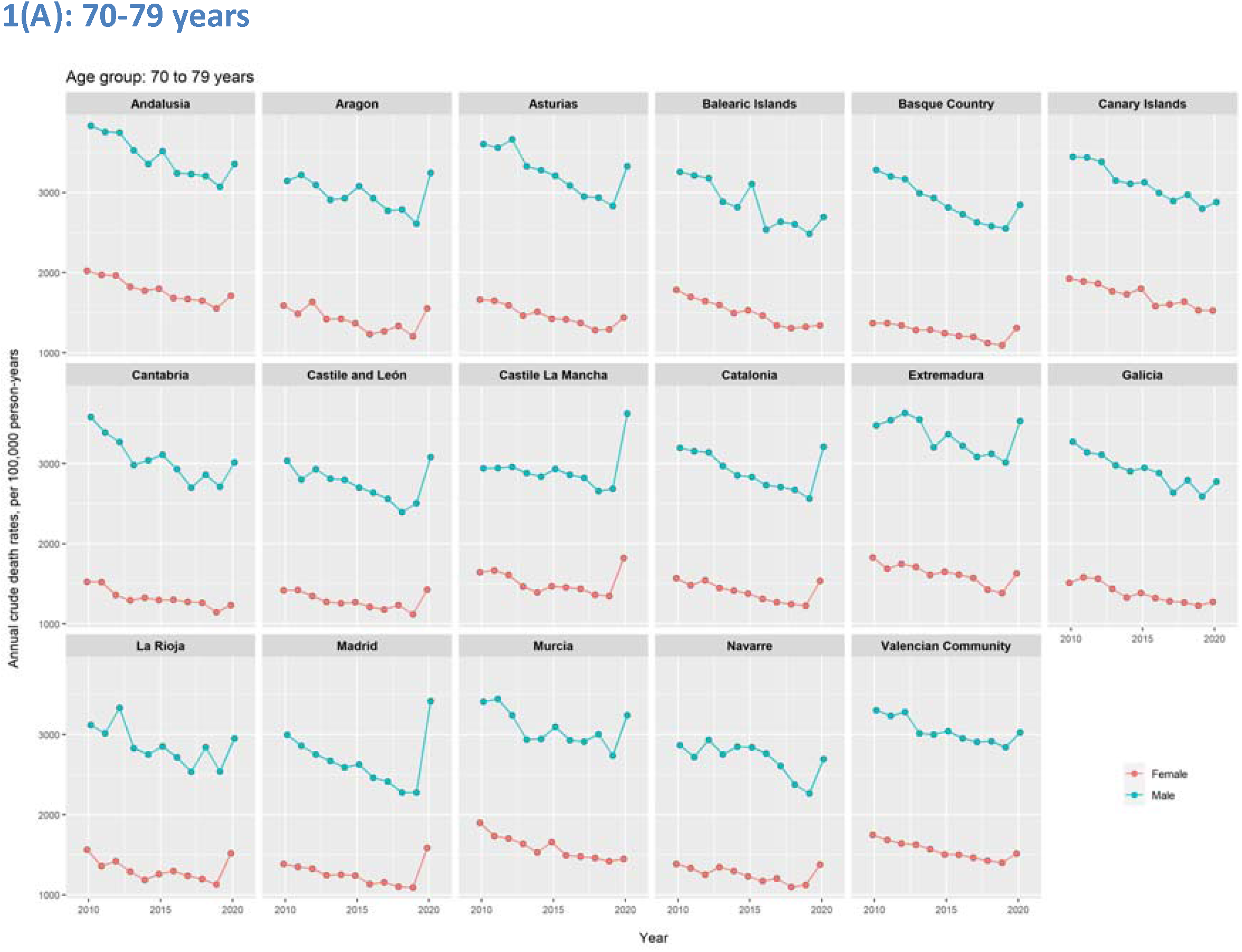
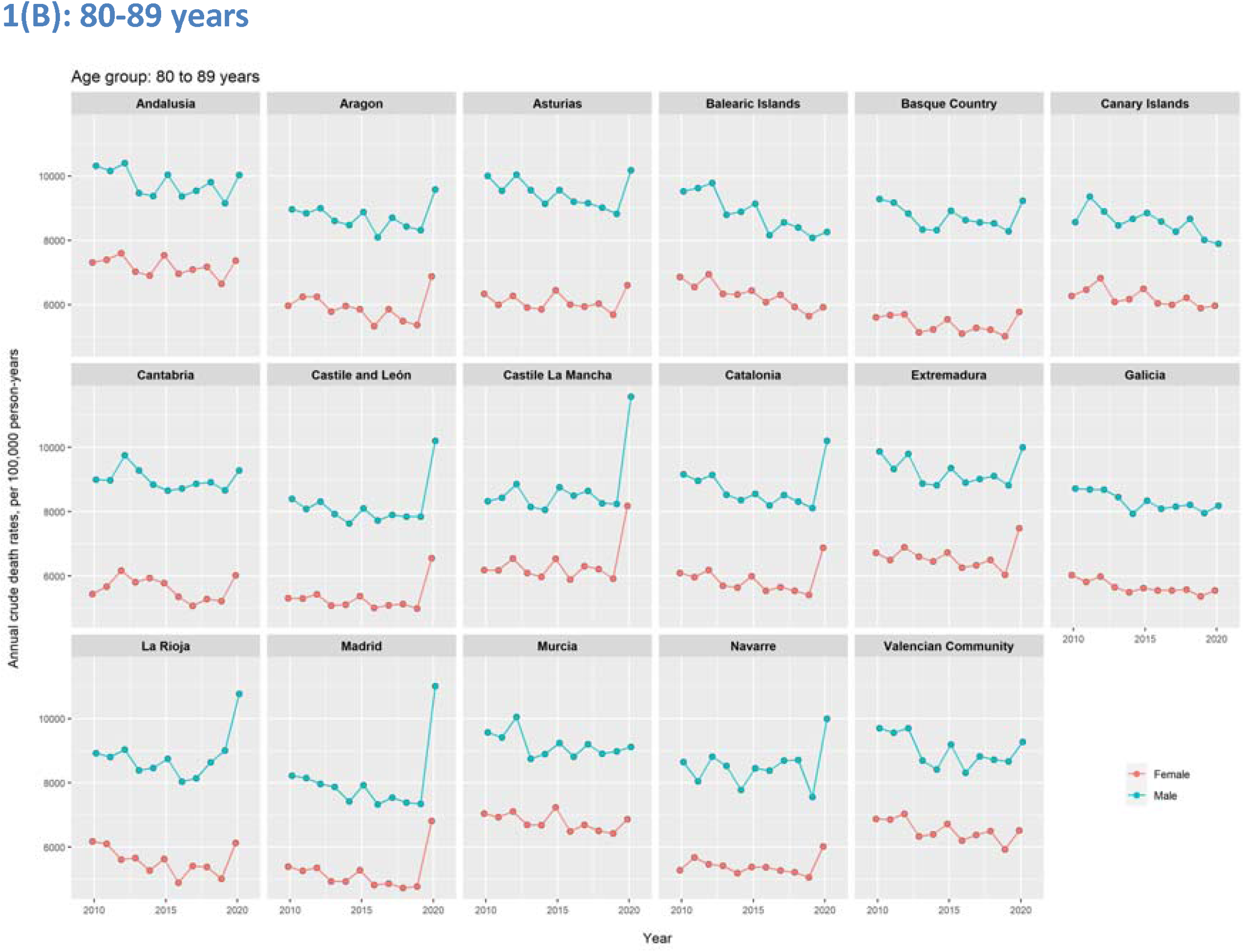
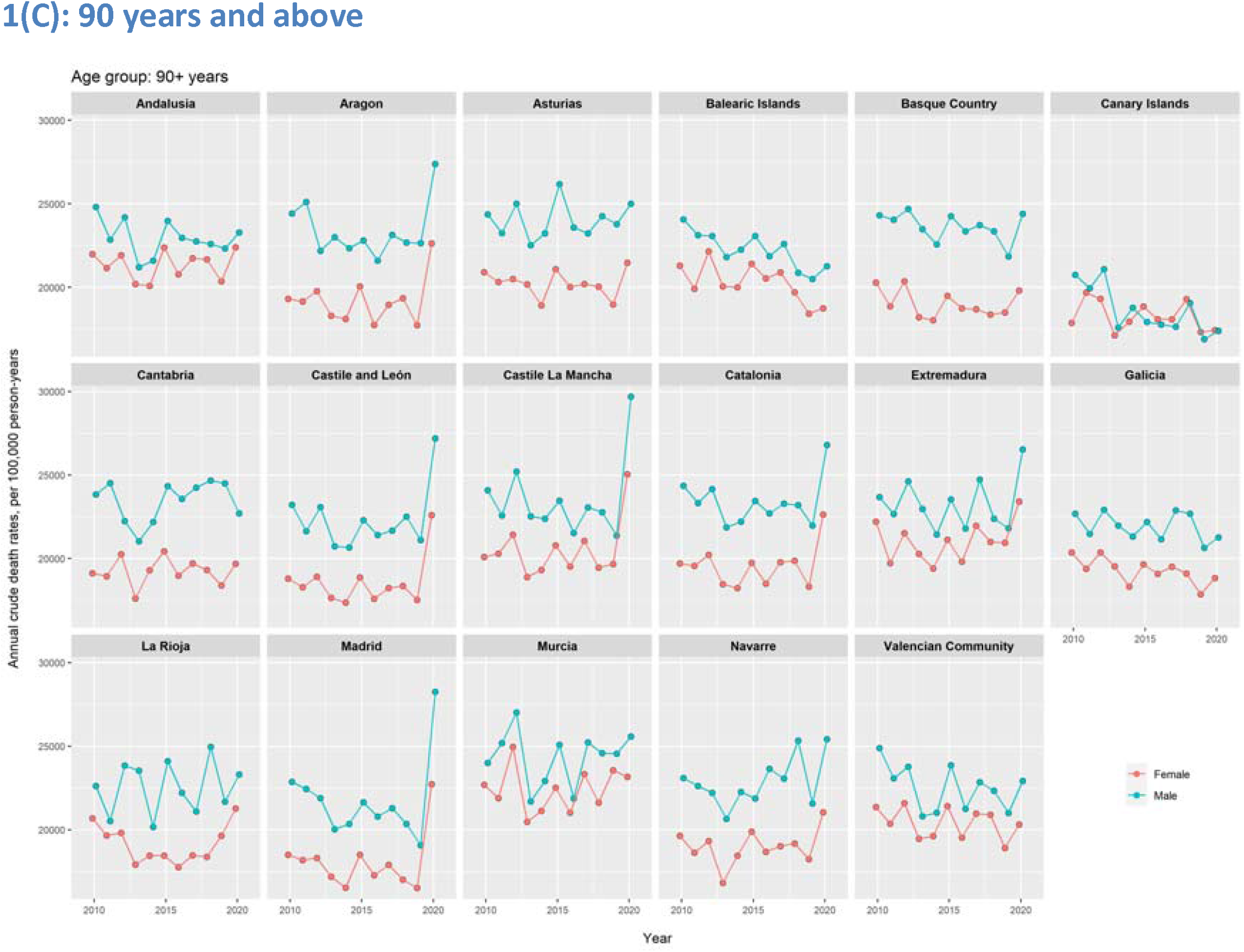
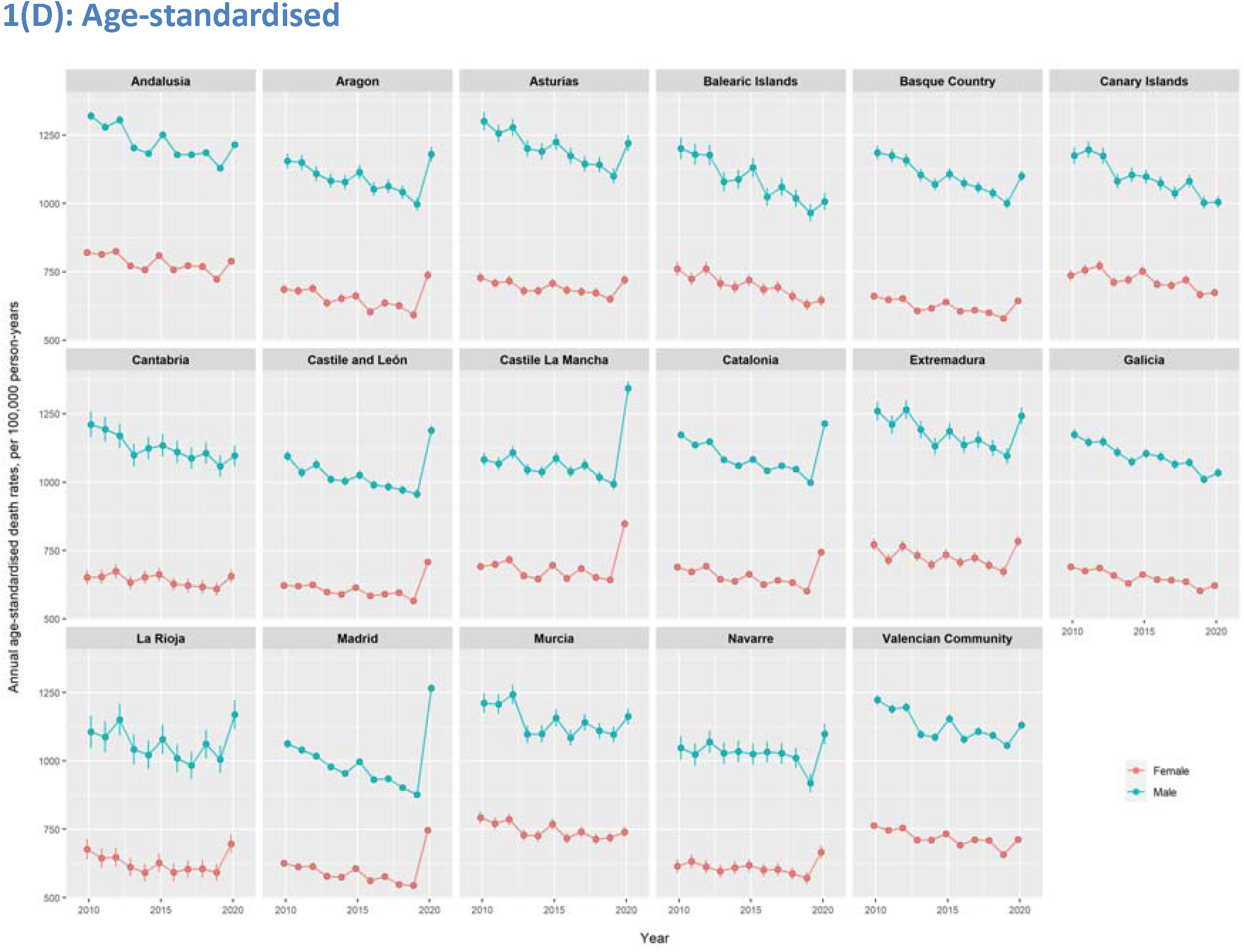
Crude and age-standardised annual mortality rate in Spanish regions, 2010-2020. -Standardised using 2013 European standard population. Confidence intervals were estimated assuming a Poisson distribution of the mortality rates. Error bar represents 95% confidence intervals.

We estimated 89,200 (95% CI: 87,600 to 90,800) excess deaths in Spain between January 2020 and June 2021. This estimate is 10% higher than the official reported Covid-19 deaths during the same period (Table 1). However, we also show a wide variability in the estimated excess deaths with the highest in Madrid (22,000; 21,500 to 22,500) and lowest in the Canary Islands (−210; -530 to 100). Compared to the reported number of Covid-19 deaths, the estimated number of excess deaths was 42% higher in Madrid. On the other hand, estimated excess deaths were lower than the reported Covid-19 deaths in several regions including Cantabria, Murcia, and the Basque Country. The shape of the epidemic curve was also highly variable across the regions and age groups, with most of the excess deaths occurring in the elderly population. The shape of the epidemic in most regions revealed two waves. Madrid and Castile La Mancha had more than 100% excess deaths during the first wave, and nearly 50% excess deaths in the second wave. Other regions that almost reached 50% excess deaths at the peak of the second wave were Aragon, and Castile and León (Figure 2 A-C).

**Table 1:** Estimated number of excess deaths in 2020-2021* in Spanish regions, by sex.

| Regions | Reported Covid-19 deaths* (A) | Percent difference between excess deaths and Covid-19 deaths | Excess deaths |  |  |
| --- | --- | --- | --- | --- | --- |
|  |  |  | Total, n (95% CI) | Men, n (95% CI) | Women, n (95% CI) |
| **Total** | **80875** | **10.2** | **89,200 (87,600 to 90,800)** | **48,000 (46,900 to 49,200)** | **41,200 (40,000 to 42,300)** |
| Madrid | 15450 | 42.3 | 22,000 (21,500 to 22,500) | 12,300 (12,000 to 12,700) | 9,700 (9,300 to 10,100) |
| Catalonia | 14736 | 27.6 | 18,800 (18,200 to 19,400) | 9,700 (9,200 to 10,100) | 9,100 (8,600 to 9,600) |
| Andalusia | 10062 | -2.6 | 9,800 (9,200 to 10,500) | 5,100 (4,700 to 5,600) | 4,700 (4,200 to 5,200) |
| Castile and León | 6925 | 2.5 | 8,100 (7,700 to 8,500) | 4,500 (4,200 to 4,800) | 3,600 (3,300 to 3,900) |
| Castile La Mancha | 6012 | 18.1 | 7,100 (6,800 to 7,500) | 4,100 (3,800 to 4,300) | 3,000 (2,800 to 3,300) |
| Valencian Community | 7412 | -11.0 | 6,600 (6,000 to 7,100) | 3,500 (3,100 to 3,800) | 3,100 (2,800 to 3,500) |
| Aragon | 3549 | 7.1 | 3,800 (3,500 to 4,100) | 1,900 (1,700 to 2,100) | 1,900 (1,700 to 2,100) |
| Basque Country | 4535 | -29.4 | 3,200 (2,800 to 3,500) | 1,700 (1,400 to 2,000) | 1,400 (1,200 to 1,700) |
| Asturias | 1978 | 31.4 | 2,600 (2,300 to 2,900) | 1,300 (1,100 to 1,500) | 1,300 (1,100 to 1,500) |
| Extremadura | 1807 | 16.2 | 2,100 (1,800 to 2,400) | 1,100 (870 to 1,200) | 1,000 (870 to 1,200) |
| Galicia | 2423 | -21.6 | 1,900 (1,500 to 2,300) | 1,100 (750 to 1,400) | 840 (530 to 1,200) |
| Navarre | 1184 | 1.3 | 1,200 (980 to 1,300) | 750 (620 to 870) | 410 (280 to 540) |
| Murcia | 1606 | -42.7 | 920 (650 to 1,200) | 570 (370 to 760) | 360 (170 to 540) |
| La Rioja | 778 | -26.7 | 570 (430 to 710) | 260 (160 to 360) | 300 (210 to 400) |
| Balearic Islands | 844 | -39.6 | 510 (290 to 740) | 270 (110 to 430) | 240 (90 to 400) |
| Cantabria | 570 | -61.0 | 220 (30 to 410) | 100 (-40 to 230) | 130 (-10 to 260) |
| Canary Islands | 791 | NA | -210 (-530 to 100) | -120 (-350 to 120) | -100 (-310 to 120) |

Excess deaths calculated as difference in observed and expected death predicted using an over-dispersed Poisson model that accounts for secular trends, and seasonal and natural variability. \*from January 2020 to June 2021;
\*\*Confirmed Covid-19 deaths reported by the autonomous regions to the National Epidemiology Centre as June 30, 2021

**Figure 2.**
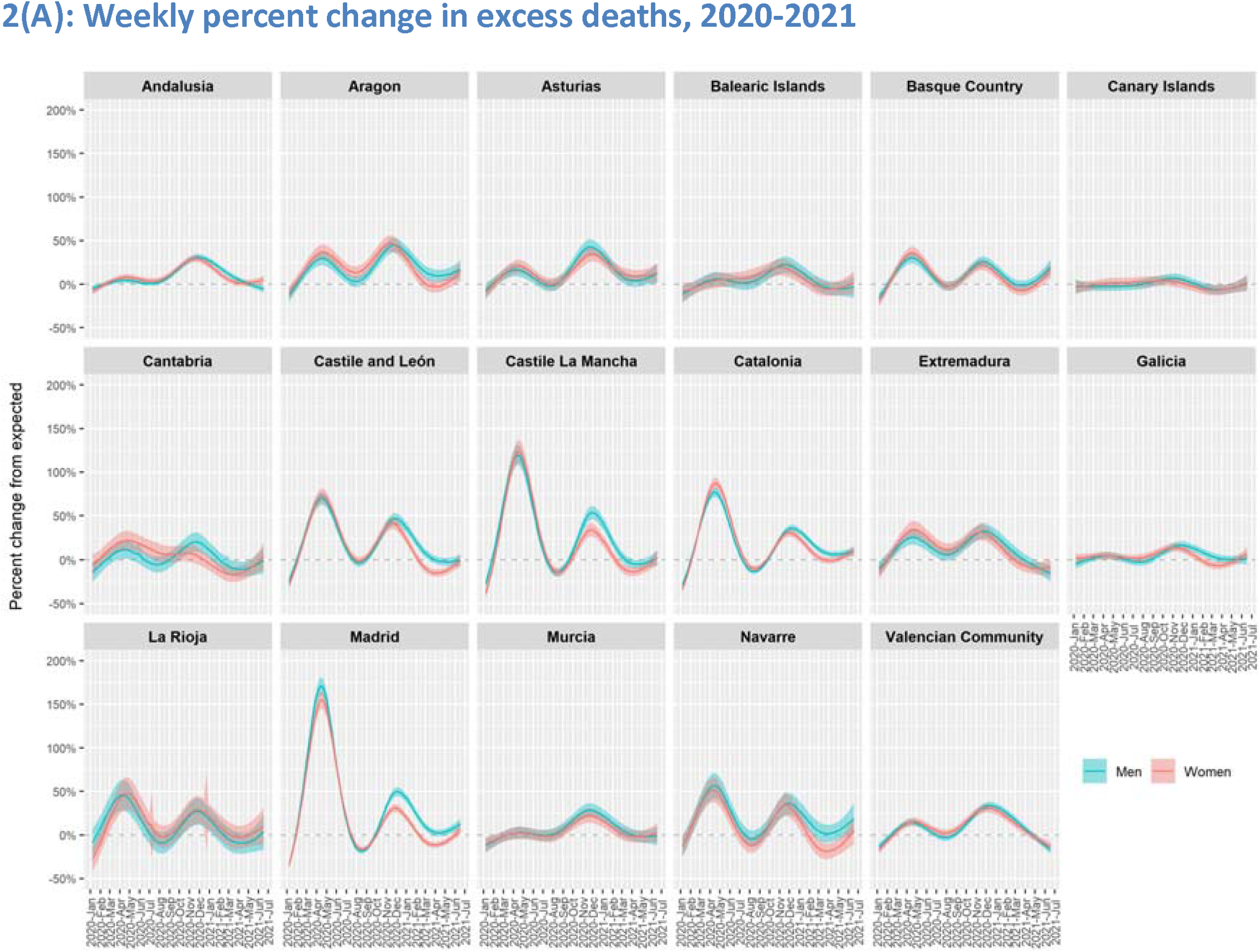
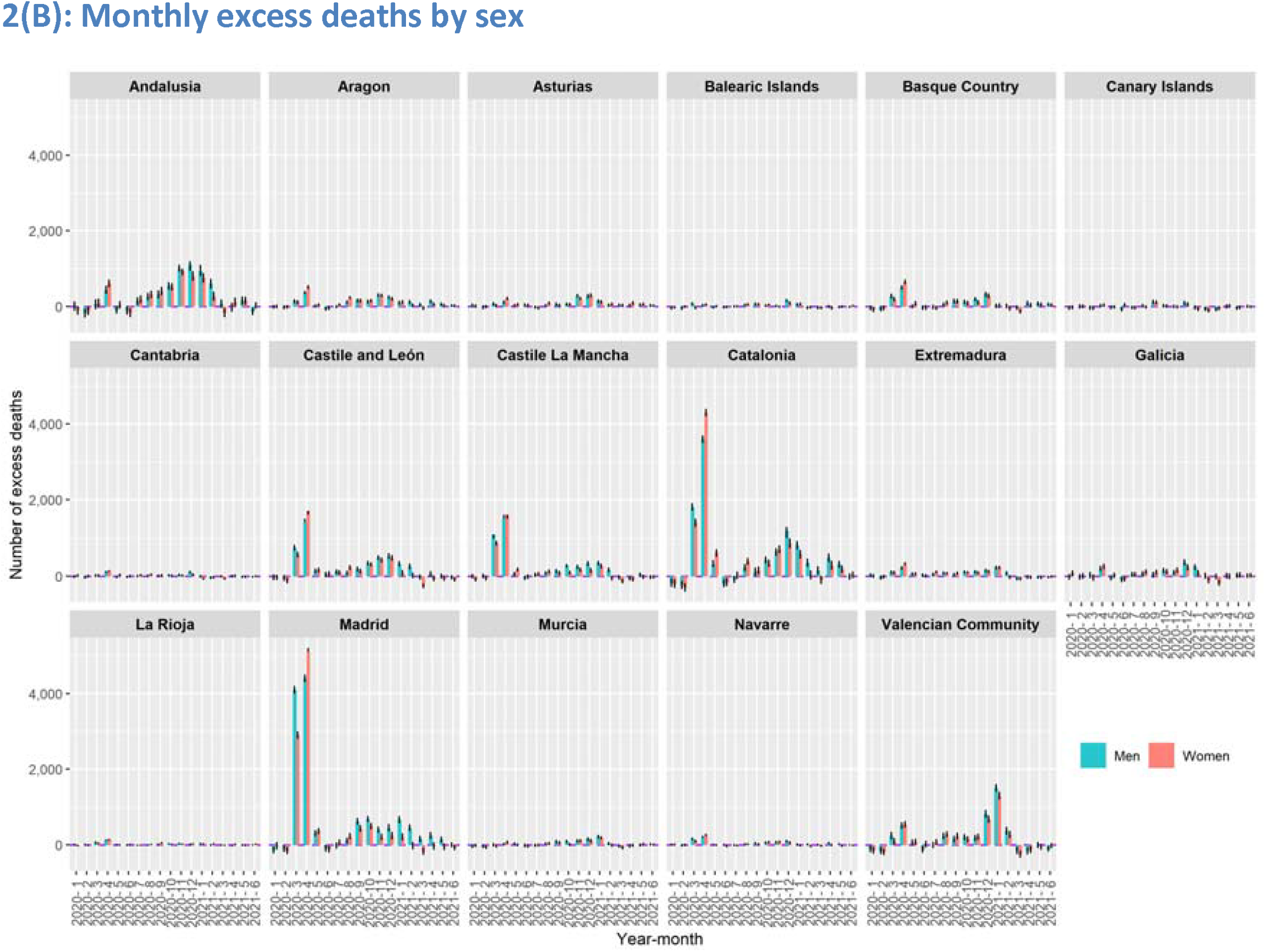
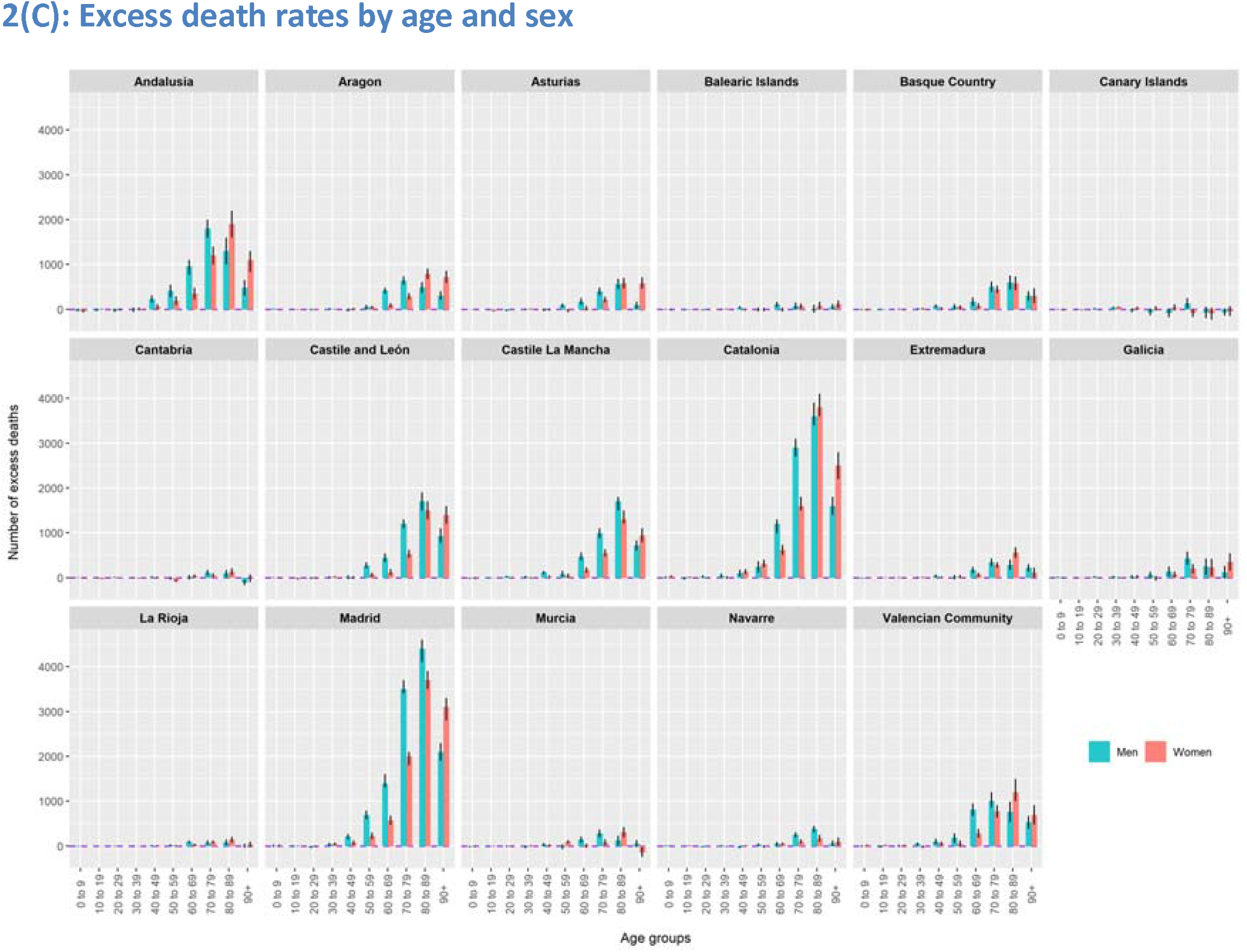
Excess deaths in Spanish regions in 2020-2021. -Standardised using 2013 European standard population. Confidence intervals were estimated assuming a Poisson distribution of the mortality rates.

Age-standardised excess death rates (per 100,000) were highest in Madrid (222.0; 216.7 to 227.4), Castile La Mancha (209.0; 198.9 to 219.0), Catalonia (155.8; 150.5 to 161.0), Aragon (155.6; 144.1 to 167.0), and Castile and León (147.4; 139.6 to 155.2). Murcia, Galicia, Balearic Islands, Cantabria, and Canary Islands had the lowest age-standardised excess death rates below 50 per 100,000. Age-standardised excess deaths were higher in men than women in most regions (Figure 3 A-C, Supplementary Table S1, and Supplementary Figure S2 A-B).

**Figure 3.**
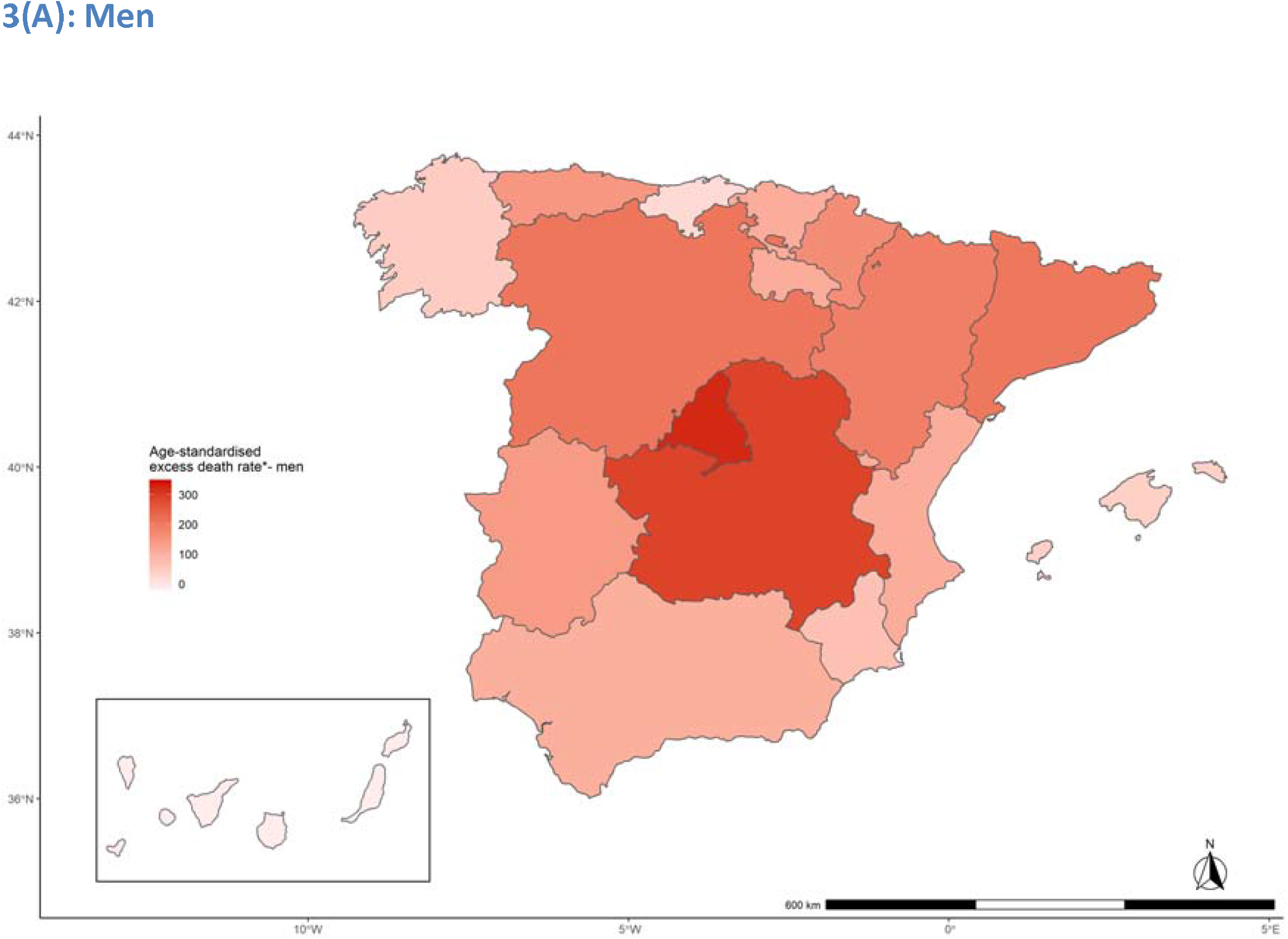
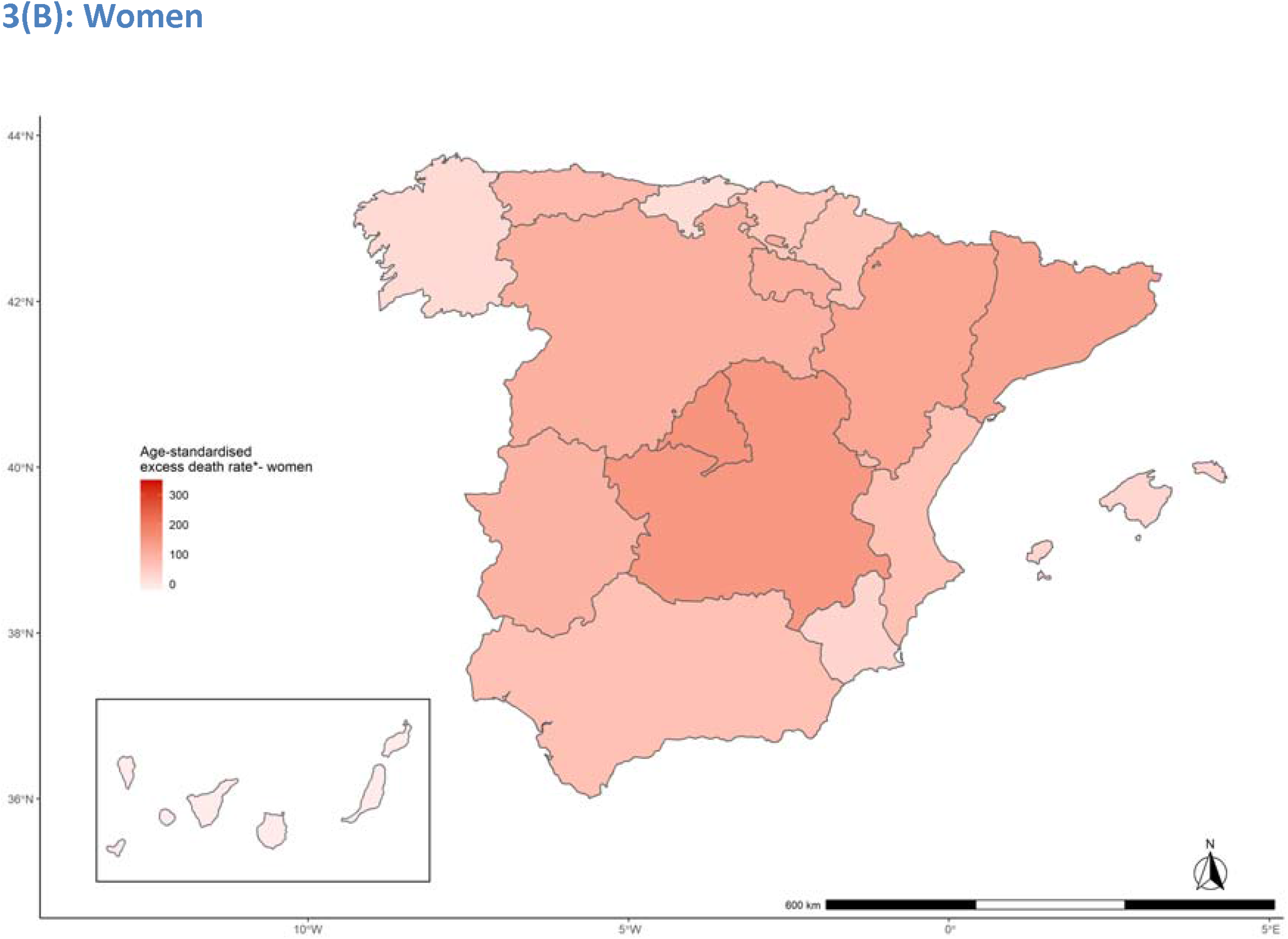
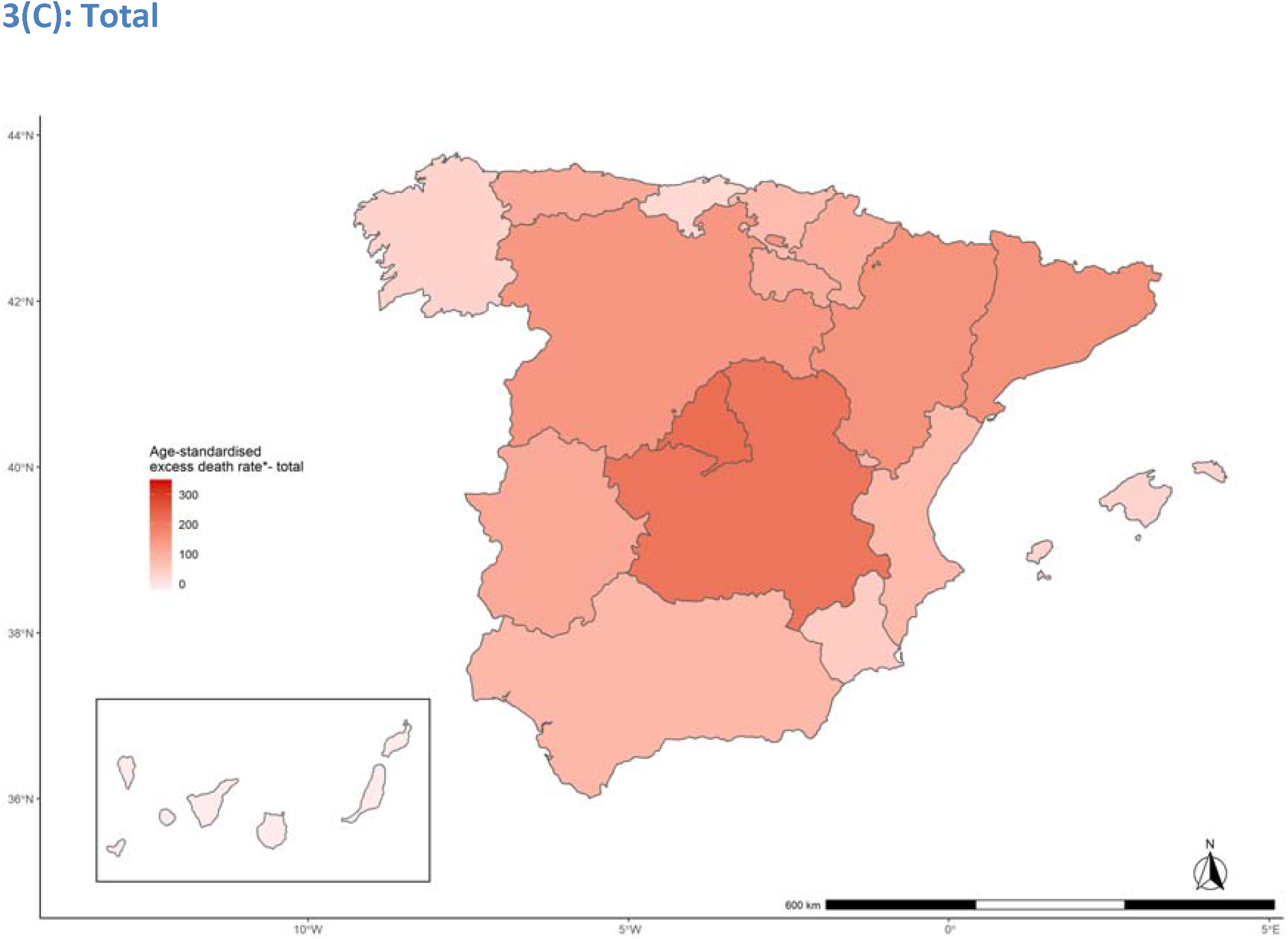
Age-standardised excess death rates in Spanish regions in 2020-2021. -Rates are per 100,000. Standardised using 2013 European standard population. Confidence intervals were estimated assuming a Poisson distribution of the mortality rates.

### Changes in life expectancy in 2020

*e*_0_ *and e*_65_ increased between 2010 and 2019 with a sharp decrease in 2020, with a steeper decline in some regions (Figure 4 A-C). Highest reduction in *e*_0_ in men was observed in Madrid (−3.48; -3.66 to -3.30), Castile La Mancha (−2.67; -3.28 to -1.62), Castile and León (−2.00; -2.13 to -1.88), Catalonia (−1.86; -2.00 to -1.71), and Aragon (−1.78; -1.94 to -1.62). Highest reduction in *e*_0_ in women was observed in Castile La Mancha (−2.30; -2.63 to -1.89), Madrid (−2.15; -2.57 to -1.72), Catalonia (−1.95; -2.17 to -1.74), Castile and León (−1.32; -1.41 to -1.23), and Aragon (−1.18; -1.62 to -0.71). Observed *e*_0_ in 2020 in men was at the level of 2006 for Castile La Mancha and 2008 for Madrid; in women, it was at the level of 2004 and 2006 for these regions, respectively.

**Figure 4.**
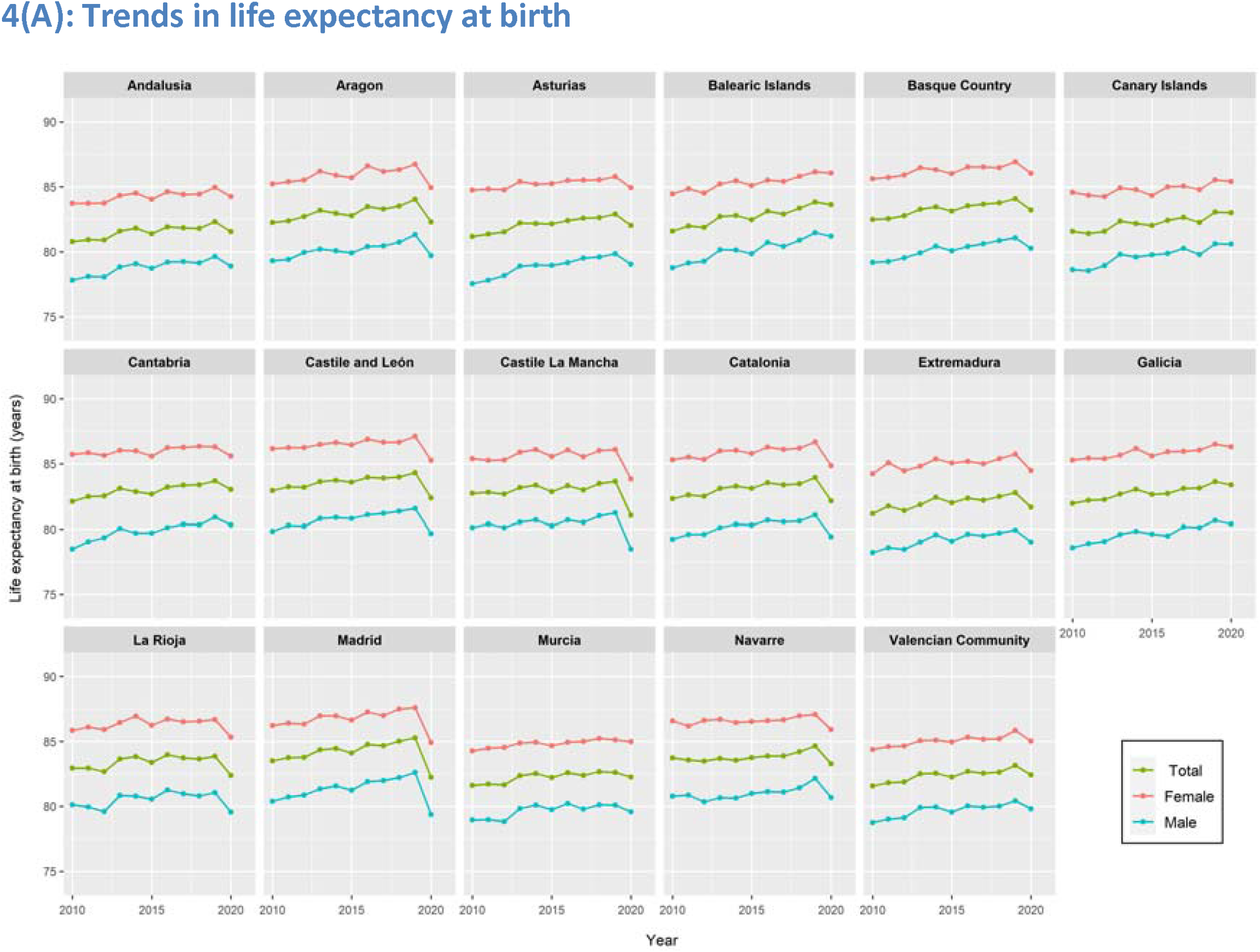
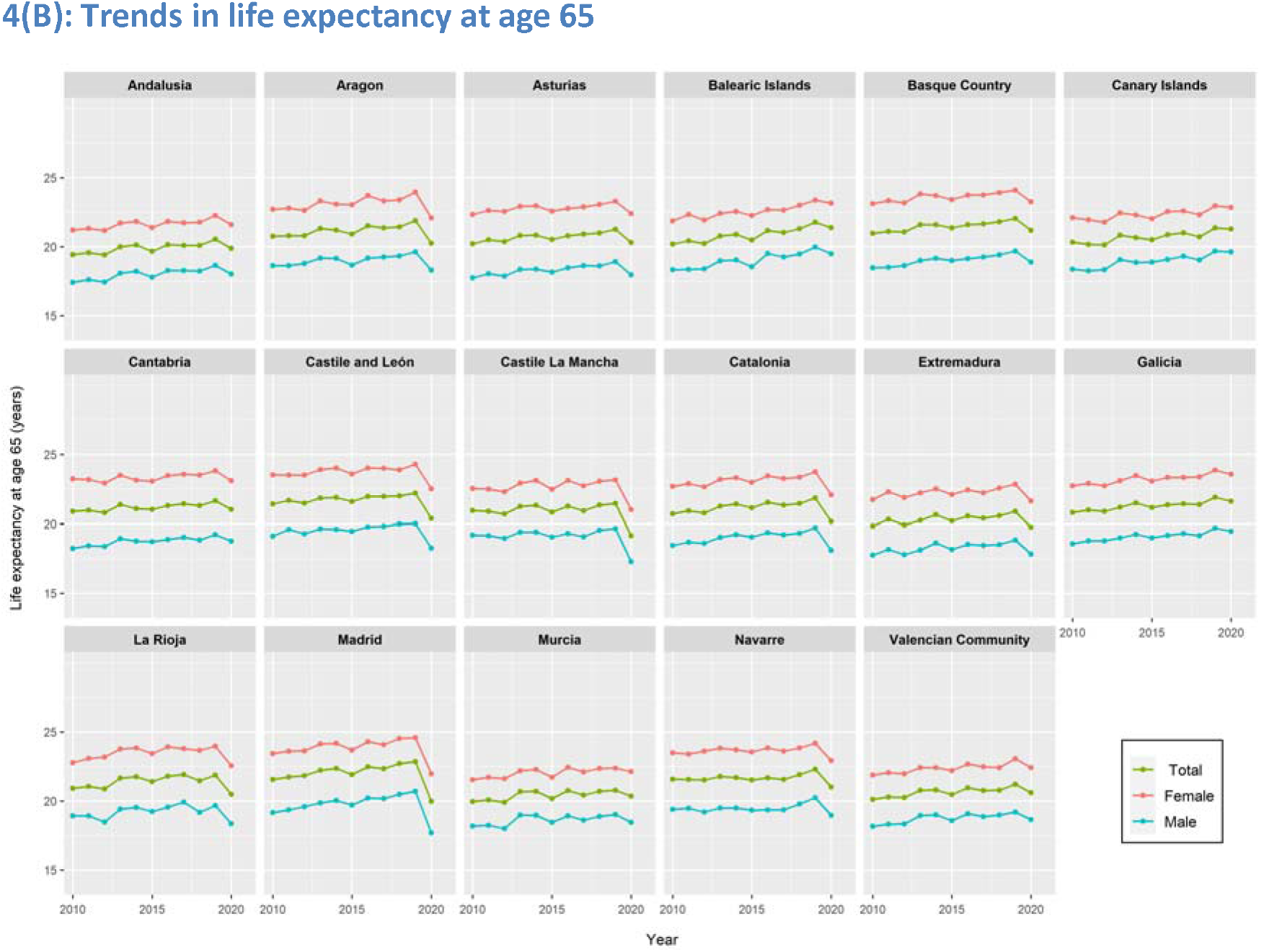
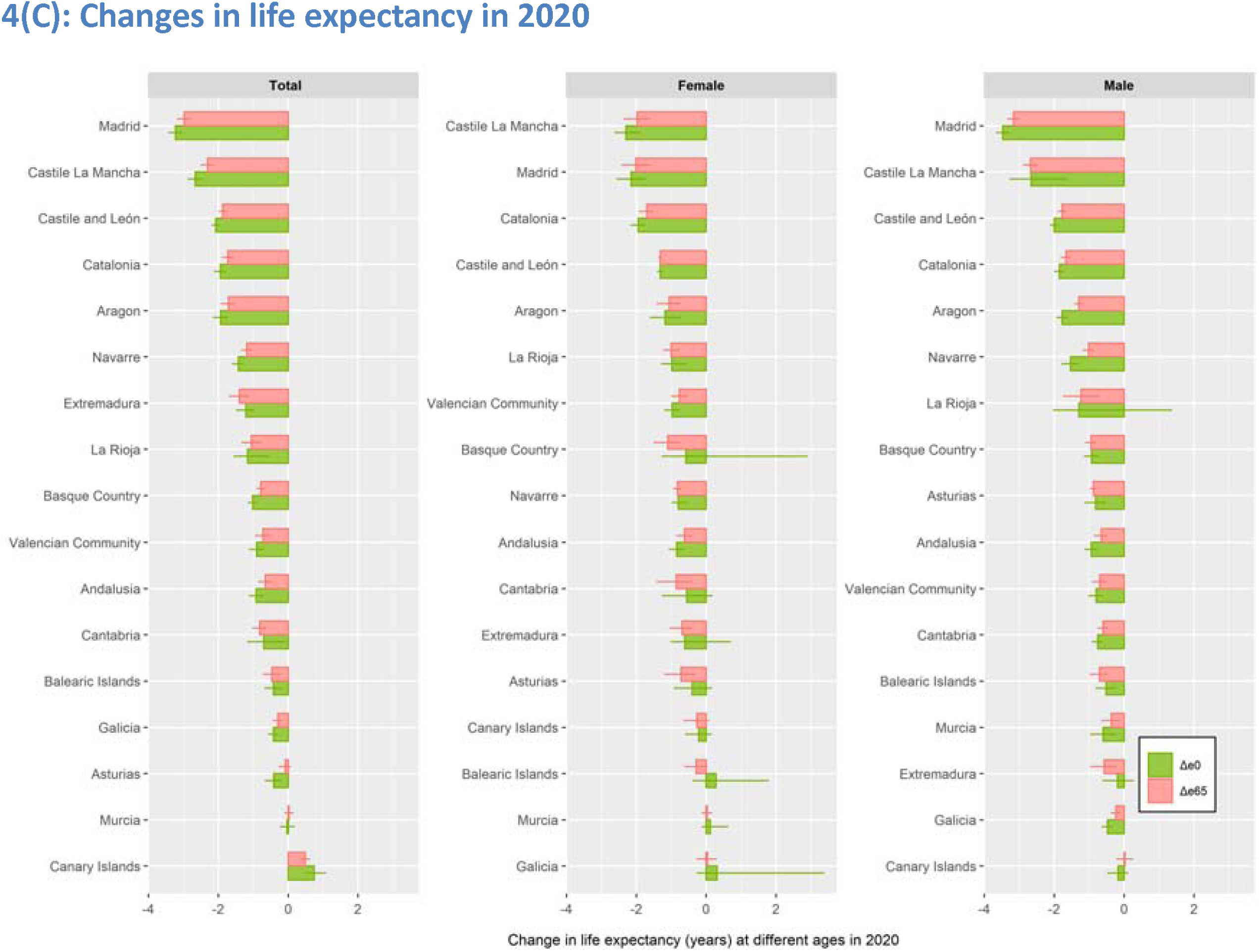
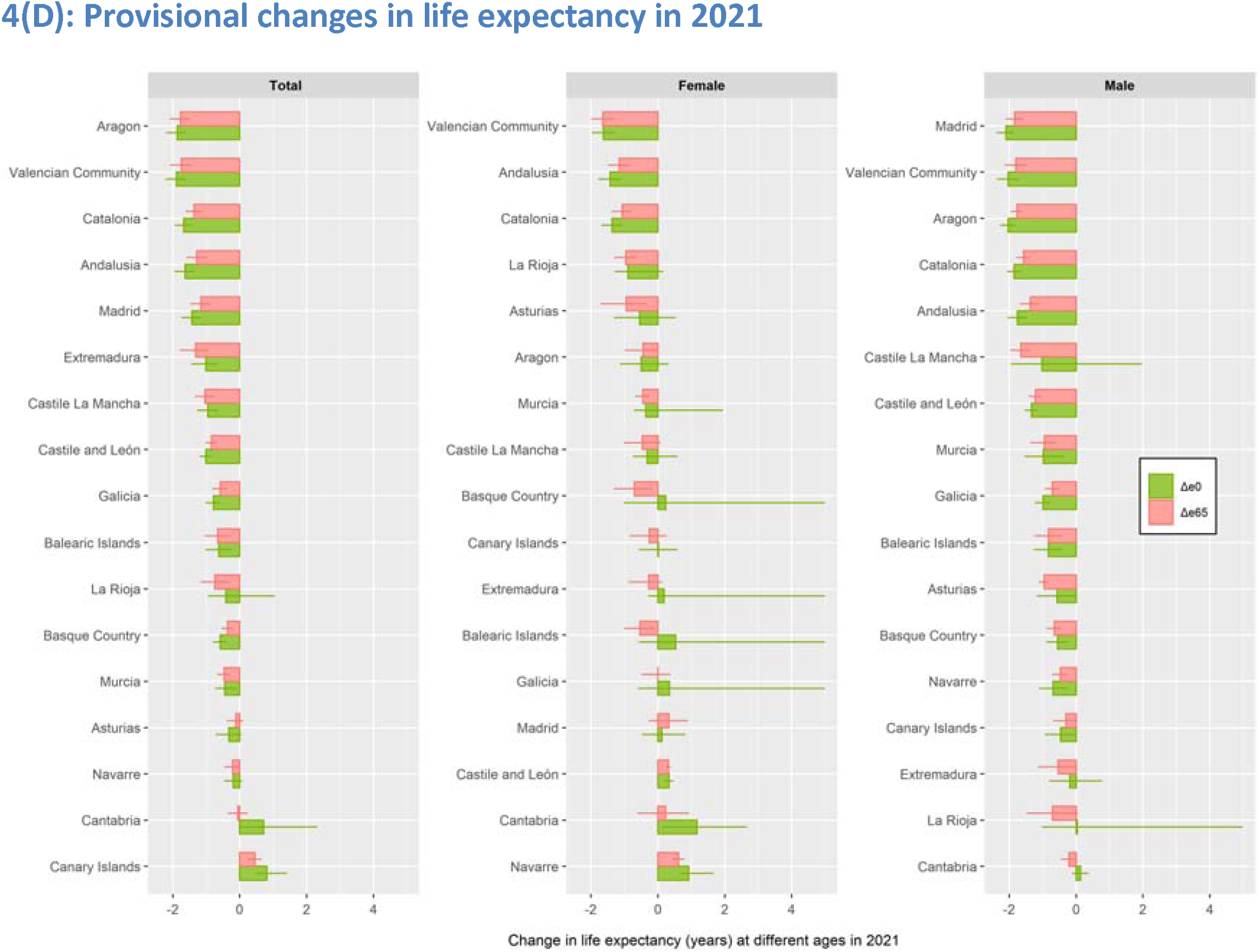
Life expectancy in Spanish regions in 2010-2021. - *Δ*e0 = changes in life expectancy at birth; *Δ*e65 = changes in life expectancy at age 65

Highest reduction in *e*_65_ in men was observed in Madrid (−3.17; -3.35 to -3.00), Castile La Mancha (−2.68; -2.89 to -2.48), Castile and León (−1.77; -1.90 to -1.65), Catalonia (−1.67; -1.81 to -1.53), and Aragon (−1.30; -1.42 to -1.18). Highest reduction in *e*_65_ in women was observed in Madrid (−2.01; -2.41 to -1.63), Castile La Mancha (−1.98; -2.35 to -1.61), Catalonia (−1.70; -1.92 to -1.49), Castile and León (−1.32; -1.37 to -1.27), and Basque Country (−1.10; -1.50 to -0.73). Observed *e*_65_ in 2020 in men in Castile La Mancha, Madrid, and Castile and León was at the level of 2000, 2004, and 2004, respectively; in women, it was at the level of 2004, 2005 and 2006 for these regions, respectively.

### Changes in YLL in 2020

YLL decreased during 2010-2019 with a sharp increase in 2020 in most regions, especially in men and elderly population (Supplementary Figure S3 A-E). Excess YLL (per 100,000) in men was highest in Castile La Mancha (5210; 3030 to 7410), Madrid (5050; 4310 to 5800), Castile and León (4100; 3410 to 4790), Aragon (3260; 2380 to 4120), and Catalonia (3050; 2310 to 3800). Excess YLL per 100,000) in women was highest in Castile La Mancha (3470; 1860 to 5080), Catalonia (2600; 1800 to 3400), Madrid (2480; 759 to 4190), Castile and León (2330; 1900 to 2770), and Aragon (1620; -465 to 3700)

### Relationship between excess deaths and excess YLL

Generally, age-standardised excess death rate was highly correlated with excess YLL (correlation coefficient: 0.91). However, there were some notable differences. For example, the age-standardised excess death rate in men was slightly lower in La Rioja (166 per 100,000) than Extremadura (179). However, excess YLL (per 100,000) was 4.3 times higher in La Rioja (2434) than Extremadura (572). Similarly, in women, Catalonia (156) had a slightly higher excess death rate than Aragon (152), but the excess YLL was more than 1.6 times higher in Catalonia (2598) than in Aragon (1618).

### Provisional changes in life expectancy in 2021

Based on data up to the 24^th^ week in 2021, *e*_0_ *and e*_65_ continued to fall in most regions (Figure 4 D). Highest reduction in *e*_0_ in 2021 in men was observed in Madrid (−2.09; - 2.37 to -1.84), the Valencian Community (−2.04; -2.36 to -1.73), Aragon (−2.03; -2.27 to - 1.79), Catalonia (−1.85; -2.07 to -1.65), and Andalusia (−1.75; -2.05 to -1.47). Highest reduction in *e*_0_ in 2021 in women was observed in the Valencian Community (−1.63; - 1.97 to -1.3), Andalusia (−1.43; -1.77 to -1.1), Catalonia (−1.37; -1.69 to -1.06), La Rioja (−0.89; -1.3 to 0.17), and Asturias (−0.55; -1.3 to -0.13). These five regions also had the highest reduction in *e*_65_ in men and women, albeit at a lower level than men.

## Discussion

Our study provides a comprehensive quantitative assessment of the impact of the Covid-19 pandemic on excess deaths, loss of life expectancy and excess years of life lost across Spanish regions. We showed that there were substantial excess deaths in all regions except the Canary Islands and Cantabria. Most excess deaths were in the central regions of Spain (Madrid, Castile La Mancha, and Castile and León), followed by Aragon and Catalonia. The age-standardised excess deaths were highest in Madrid, Castile La Mancha, Catalonia, Aragon, and Castile and León. While the estimated number of excess deaths was 40% higher than the official reported Covid-19 deaths during the same period in Madrid, excess deaths were lower than the reported Covid-19 deaths in eight out of the seventeen regions. Despite a recent trend of declining annual mortality rate between 2016 and 2019, that mortality rate increased in 2020 in most regions in both men and women, more sharply for those over 60 years of age. Highest reduction in *e*_0_ in men was observed in Madrid, Castile La Mancha, Castile and León, Catalonia, and Aragon. The observed reduction in *e*_0_ and *e*_65_ in 2020 set back the life expectancy as far as 20 years in some regions.

To our knowledge, this is the most detailed and comprehensive analysis of the impact of the Covid-19 pandemic in Spanish regions using three important and complementary parameters of excess and premature mortality. Our analytic approach accounted for recent improvements in mortality and seasonal trends by age and sex in each of these regions. Several previous estimates of excess deaths have been published. The first one, reported in a press release by the National Statistics Office, which compared total mortality in 2020 with 2019, estimated the annual variation rate of the number of deaths for 2020, with 17.7% excess deaths for the whole country and 5 regions exceeding 20% excess deaths (41.2% in Madrid, 32.3% in Castile La Mancha, 26.0% in Castile and León, 23.5% in Catalonia, and 22.5% in Aragon).[41] The second analysis used the 101,938 excess deaths reported by the National Statistics Office from January 2020 to February 2021 and compared the difference between the observed mortality in that period with the expected deaths, assuming those excess deaths had been distributed proportionally to the size of the population in each region. Madrid and its two nearby areas of influence, Castile-La Mancha, and Castile and León, had the highest excess mortality (48%, 74% and 80%, respectively) and the Canary Islands and other eight regions had lower deaths than expected.[42] Moreover, García-García et al.[43] published a paper reporting excess deaths in Spain from 3 March to 29 November 2020 using the Mortality Monitoring (MoMo) surveillance system, designed to assess peaks when observed deaths from any cause are higher than those expected—as the average of the last 10 years—for at least two consecutive days.[44] Crude excess death rates per 100,000 population were higher in Castile La Mancha (347), Madrid (261), and Castile and León (244) whereas the Canary Island had the lowest rate (43). Finally, the European Committee on the Regions in a report of excess deaths in 2020 compared with the average number of deaths between 2016 and 2019, showed that Madrid was the hardest hit region in Europe, 44%, followed by Lombardia, Italy, 39%, and Castile La Mancha, 34%.[45] However, this report did not take into account the recent improvements in mortality and seasonal trends by age and sex in each European region.

We previously reported a national decline in life expectancy of −1.35 (−1.72 to −0.99) years in Spain in 2020.[26] However, this analysis shows large variations in life expectancy loss across regions. Using partial data up to July 5, 2020, a previous study reported a reduction in life expectancy with the highest reduction of 2.8 years in Madrid.[27] This study compared the 2020 provisional estimates with 2019 estimates, which may have contributed to an underestimation of the effect on life expectancy across all the regions.[25] Comparing the 2020 estimates with those in 2019 or an average of the most recent few years may lead to incorrect conclusions because it ignores recent trends in mortality.[25] We are not aware of any previous studies that have estimated the effects of the pandemic on YLL in Spanish regions.

Our findings have important policy implication. Reported Covid-19 deaths were lower than the number of excess deaths in nine of the regions included in our study. This suggests a possible underestimation and/or incomplete identification and reporting of the Covid-19 deaths, as well as the deaths that would have not happened had the Covid-19 pandemic not occurred, due to the health system malfunction and/or collapse in some regions. In contrast, the number of reported Covid-19 deaths were smaller than the number of excess deaths in the remaining eight regions. For instance, in the Basque Country, 3200 excess deaths were estimated by the model while 4535 confirmed Covid-19 deaths were registered. Therefore, there were 1335 deaths fewer than expected, despite the reported Covid-19 deaths. Those could have been the deaths that would have happened had the Covid-19 pandemic not occurred. This is an example of ‘avoided mortality’, which were prevented by behavioural and environmental changes due to the Covid-19 pandemic, such as wearing a facemask, or decreased air pollution, fewer occupational and traffic injuries, as was seen in 2008 economic crisis.[46] The regions that may have had fewer deaths than expected for causes other than Covid-19 were the Valencian Community, Basque Country, Galicia, Murcia, La Rioja, Balearic Islands and Cantabria and, to a lesser extent, Andalusia. In most of these regions, the epidemic in Spring 2020 was less severe and health care services did not collapse, as was the case for regions with a strong epidemic in Spring 2020, particularly in Madrid and Castile La Mancha, the only two regions where weekly excess deaths reached more than 100% during the first wave. We hypothesize that efforts to prevent the spread of Covid-19 and associated environmental changes resulted in the prevention of deaths from respiratory diseases and occupational injuries that would have occurred had the Covid-19 epidemic not occurred.[47]

It is worth noting that the observed number deaths in Canary Islands was lower than expected despite having 791 reported Covid-19 deaths. This might have been related to the fact that the Canary Islands quickly responded at the start of the pandemic. The Islands reported its first case of Covid-19, a German tourist at La Gomera, on 31 January 2020, which was arguably the first case of Covid-19 in Spain. The local authority responded quickly with isolation of the detected case and contact tracing, and thereby, avoided an early spread of the pandemic in the Canary Islands. After the second case was diagnosed, public health authorities in the Canary Islands applied strict quarantine measures to prevent SARS-CoV-2 from spreading. In Madrid, the region with the highest excess deaths, the first case was diagnosed on 25 February 2020, although retrospective investigation suggested that Madrid probably had the first Covid-19 case diagnosed in Spain, by early January 2020,[43] and the counts grow exponentially in late February and early Mach, according to the official information at *https://cnecovid.isciii.es/covid19/*. By the time public health authorities implemented measures in Madrid, in early March 2020, widespread community transmission of SARS-COV-2 was happening.

Important differences in resources employed and measures applied to contain SARS-CoV-2 spreading may have had accounted for much of the differences in excess deaths and reduction in life expectancy across the regions. In this respect, Madrid had an ill-prepared and underfunded public health system compared to most other regions. For example, 2018 data show that public spending on health, the percentage of public expenditure on primary care, and the rate of primary care nurses in Madrid were the lowest of all regions, 3.6%, 11.0% and 0.5 per 1000 inhabitants against an average of 5.5%, 13.9% and 0.7 per 1000 inhabitants, respectively.[48] Madrid was also the region that applied the least stringent measures to contain the spread of Covid-19 infection.[49–51]

The variability in excess deaths found in the regions of Spain during the first wave is consistent with the variability found in the first national seroprevalence study of SARS-CoV-2 (ENE-COVID Study) conducted in the community-dwelling population.[52] According to this survey conducted between April 27 and May 11 of 2020, Madrid, the neighbouring five provinces in Castile La Mancha and two close provinces of Castile and León had the highest prevalence of IgG antibodies to SARS-Cov-2. To a certain extent, excess deaths and high seroprevalence of SARS-Cov-2 antibodies in provinces close to Madrid could have been related to population mobility, in the absence of measures to contain the spread of the virus. According to a 2019 survey conducted by the National Statistics Institute, more than 160,000 people were travelling daily to work in Madrid from other provinces, and about 120,000 came from four neighbouring provinces in Castile-la Mancha and Castile and León.[53]

In the latest round of the national survey, conducted in November 2020,[54] the region of Madrid continued to have the highest prevalence (12.4%) of SARS-CoV-2. Neighbouring four provinces of Castile La Mancha, and six provinces of Castile and León had prevalence rates higher than 13%. Only one additional province, Navarra, had such a high SARS-CoV-2 prevalence rate out of the remaining 41 provinces or autonomous cities of Spain. Although the ENE-COVID study was limited to the community-dwelling population and specifically excluded institutionalized populations, excess deaths highly reflected the prevalence rate of Covid-19 infection in the community.

In Spain, as in many high-income countries, Covid-19 deaths have occurred primarily in nursing homes. Older adults living in nursing homes constitute less than 1% of the total population, and less than 4% of those aged 65. However, they have contributed 30,507 of the total 82,125 covid-19 deaths (37.1%) officially reported since the beginning of the pandemic until August 8, 2021. Some protocols were applied in several regions of Spain during the spring of 2020 regarding their healthcare. More specifically, the Madrid Government issued orders to prohibit the referral to public hospitals of nursing home residents with severe disability or cognitive impairment. As a result, nursing home residents did not receive adequate medical management; many residents died alone in the nursing homes.[55,56]

This study confirms that, in Spain, the existing sex inequality in the annual standardised mortality rate, with higher rates in men than in women, was further widened in almost all regions during the study period, in agreement with what has been observed in a previous study on 29 high income countries during the Covid-19 pandemic.[22]

The analyses presented here are based on newly developed methods that allow the examination of excess deaths and premature mortality by age groups and sex. The analysis also considers period and seasonal changes that have proved to be sensitive to environmental and societal changes. This methodology has advantages over alternatives, such as those used to assess the excess deaths associated with the macroeconomic changes in Spain during the Great Recession.[46,57] The accuracy of the number of deaths and population by region, age group and sex provided by the National Statistics Office is robust and reliable, although the most recent figures may have some small errors. The estimates for the changes in life expectancy might be slightly biased because they are based on partial data for the first six months, which in normal circumstances have a slightly higher mortality than the second half of each year.

Our study provides new evidence on the direct and indirect effects of the Covid-19 pandemic on excess and premature mortality. It underscores the importance of the availability of age and sex disaggregated data for more nuanced analysis, and estimation of the direct and indirect effects of the pandemic. The Canary Islands and Cantabria stood out as the two regions that had a lower-than-expected mortality across all the age groups, in both men and women, which could potentially be attributed to the country’s prevention and control strategy early in the pandemic. Our findings also suggest that many regions had an underestimation or underreporting of Covid-19 deaths, a substantial increase in non-covid-19 deaths, or both.

This study also highlights the potential value of more reliable and timely monitoring of excess deaths at both a national and regional levels, to inform public health policy to mitigate the impact of the pandemic on excess deaths and premature mortality. Furthermore, such data would help to detect important social inequalities in the impact of the pandemic and inform more targeted interventions. A concerted effort to improve the sharing of data, and knowledge on the impact of public health policy, would improve the present response to the pandemic in Spain and its regions, and would improve the resilience of the country to future pandemics.

## Data Availability

Annual population data, and the weekly mortality data from first week of 2019 and to week 24 of 2021 are publicly available from the Instituto Nacional de Estadistica (https://www.ine.es/jaxiT3). We have obtained weekly data for 2015-2018 (for excess mortality), and monthly data (including infant mortality) for 2010-2020 (for life expectancy and years of life lost calculation) from Instituto Nacional de Estadistica with a small fee. These data could be obtained from Instituto Nacional de Estadistica (with a small fee), but we will make the data available on a public repository if this is approved by the Instituto Nacional de Estadistica.

