## Supplementary for "Unequal impact of the Covid-19 pandemic on excess deaths, life expectancy, and premature mortality across Spanish regions in 2020 and 2021"

Supplementary Materials

Supplementary Table 1: Crude and age-standardised excess death rate per 100,000 in 2020-2021*

| | Excess death rate per 100,000 person-years, crude | | | Excess death rate per 100,000 person-years, age-standardised | | |
|---|---|---|---|---|---|---|
| Regions | Total, n (95% CI) | Men, n (95% CI) | Women, n (95% CI) | Total, n (95% CI) | Men, n (95% CI) | Women, n (95% CI) |
| Madrid | 222.8 (217.7 to 227.9) | 259.9 (253.6 to 268.4) | 188.6 (180.9 to 196.4) | 222.0 (216.7 to 227.4) | 325.4 (315.5 to 335.2) | 154.0 (147.9 to 160.1) |
| Castile La Mancha | 237.8 (227.7 to 251.2) | 273.5 (253.5 to 286.9) | 201.7 (188.3 to 221.9) | 209.0 (198.9 to 219.0) | 291.3 (274.2 to 308.4) | 146.1 (134.3 to 158.0) |
| Catalonia | 168 (162.6 to 173.4) | 177.2 (168.1 to 184.5) | 159.2 (150.5 to 168) | 155.8 (150.5 to 161.0) | 204.8 (195.4 to 214.3) | 121.2 (115.2 to 127.2) |
| Aragon | 195.3 (179.9 to 210.7) | 197.8 (177 to 218.6) | 192.9 (172.6 to 213.2) | 155.6 (144.1 to 167.0) | 192.6 (172.4 to 212.7) | 119.9 (106.7 to 133.2) |
| Castile and León | 231.7 (220.3 to 243.2) | 260.9 (243.5 to 278.3) | 203.4 (186.4 to 220.3) | 147.4 (139.6 to 155.2) | 207.7 (194.2 to 221.3) | 101.3 (92.3 to 110.3) |
| Extremadura | 135.6 (116.3 to 155) | 143.2 (113.3 to 156.2) | 128.2 (111.5 to 153.8) | 114.3 (100.3 to 128.2) | 140.5 (116.1 to 164.9) | 94.9 (79 to 110.9) |
| Asturias | 175.2 (155 to 195.4) | 183.3 (155.1 to 211.5) | 167.8 (142 to 193.6) | 112.2 (99.4 to 124.9) | 147.7 (124.3 to 171) | 82.8 (68.2 to 97.5) |
| Navarre | 124.9 (102 to 135.3) | 158.1 (130.7 to 183.3) | 84.4 (57.6 to 111.1) | 106.4 (90.1 to 122.8) | 166.9 (137.7 to 196.1) | 62.8 (43.8 to 81.7) |
| La Rioja | 123.4 (93.1 to 153.7) | 114.2 (70.3 to 158.2) | 128 (89.6 to 170.6) | 105.5 (81.5 to 129.6) | 109.1 (66.1 to 152.1) | 100 (72.8 to 127.3) |
| Andalusia | 79 (74.2 to 84.7) | 83.4 (76.8 to 91.5) | 74.8 (66.9 to 82.8) | 85.6 (79.7 to 91.4) | 103.4 (93.2 to 113.6) | 69.4 (62.6 to 76.3) |
| Valencian Community | 89.7 (81.5 to 96.4) | 96.6 (85.5 to 104.8) | 83 (74.9 to 93.7) | 84.7 (77.9 to 91.5) | 105.5 (93.6 to 117.4) | 67.3 (59.4 to 75.2) |
| Basque Country | 100 (87.5 to 109.4) | 109.8 (90.4 to 129.2) | 84.8 (72.7 to 102.9) | 77.9 (69.1 to 86.6) | 108.4 (92.2 to 124.5) | 59.4 (49.5 to 69.3) |
| Murcia | 41.7 (29.5 to 54.4) | 51.6 (33.5 to 68.9) | 32.7 (15.4 to 49) | 47.2 (33.3 to 61.2) | 71.3 (46.9 to 95.6) | 30.9 (14.6 to 47.2) |
| Galicia | 48.2 (38 to 58.3) | 57.8 (39.4 to 73.6) | 41.2 (26 to 58.8) | 34.4 (26.8 to 42.1) | 48.8 (35.1 to 62.4) | 23 (14.3 to 31.6) |
| Balearic Islands | 28.7 (16.3 to 41.7) | 30.4 (12.4 to 48.4) | 27 (10.1 to 45.1) | 33 (18.5 to 47.5) | 41 (15.5 to 66.5) | 28.4 (11.4 to 45.3) |
| Cantabria | 25.8 (3.5 to 48.2) | 24.1 (-9.6 to 55.5) | 29.8 (-2.3 to 59.6) | 23.9 (6.4 to 41.4) | 22.9 (-9.5 to 55.2) | 16 (-3.9 to 35.9) |
| Canary Islands | -6.4 (-16.2 to 3) | -7.4 (-21.5 to 7.4) | -6 (-18.7 to 7.2) | -7.8 (-18.9 to 3.2) | -9.6 (-28.3 to 9.2) | -7.7 (-20.9 to 5.5) |

Excess deaths calculated as difference in observed and expected death predicted using an over-dispersed Poisson model that accounts for secular trends, and seasonal and natural variability. *from January 2020 to June 2021.

Supplementary Figure S1: Annual mortality rate in Spanish regions, 2010-2020

S1 (A): <10 years

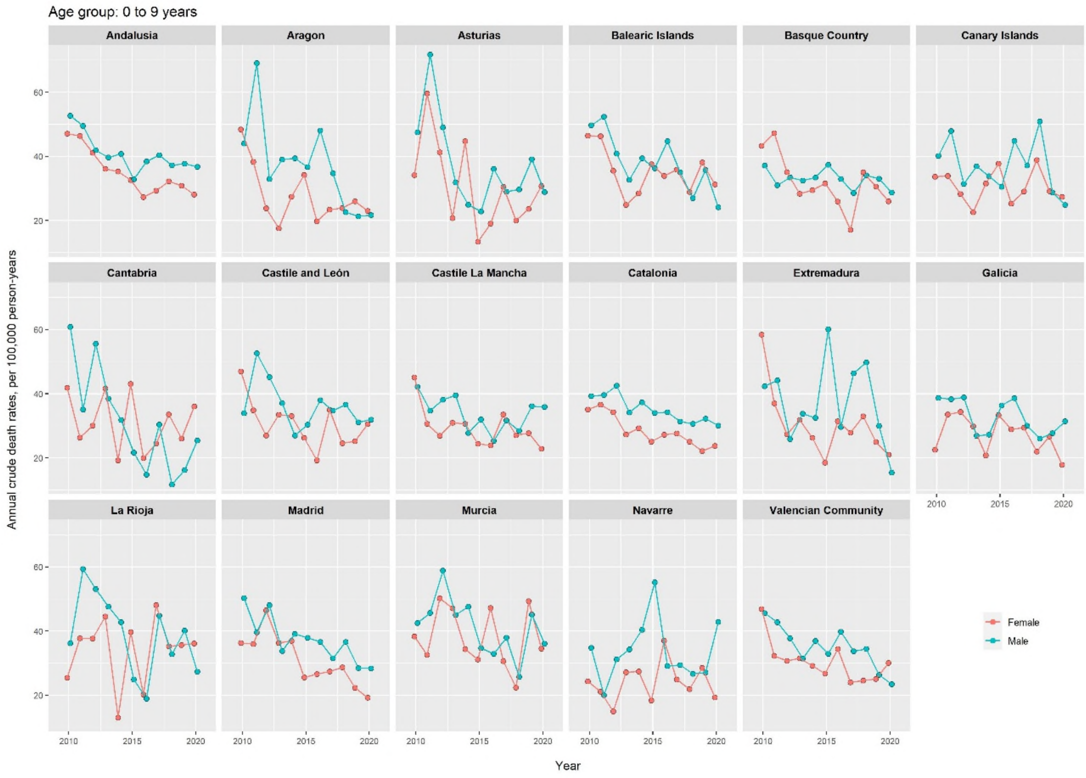

S1 (B): 10-19 years

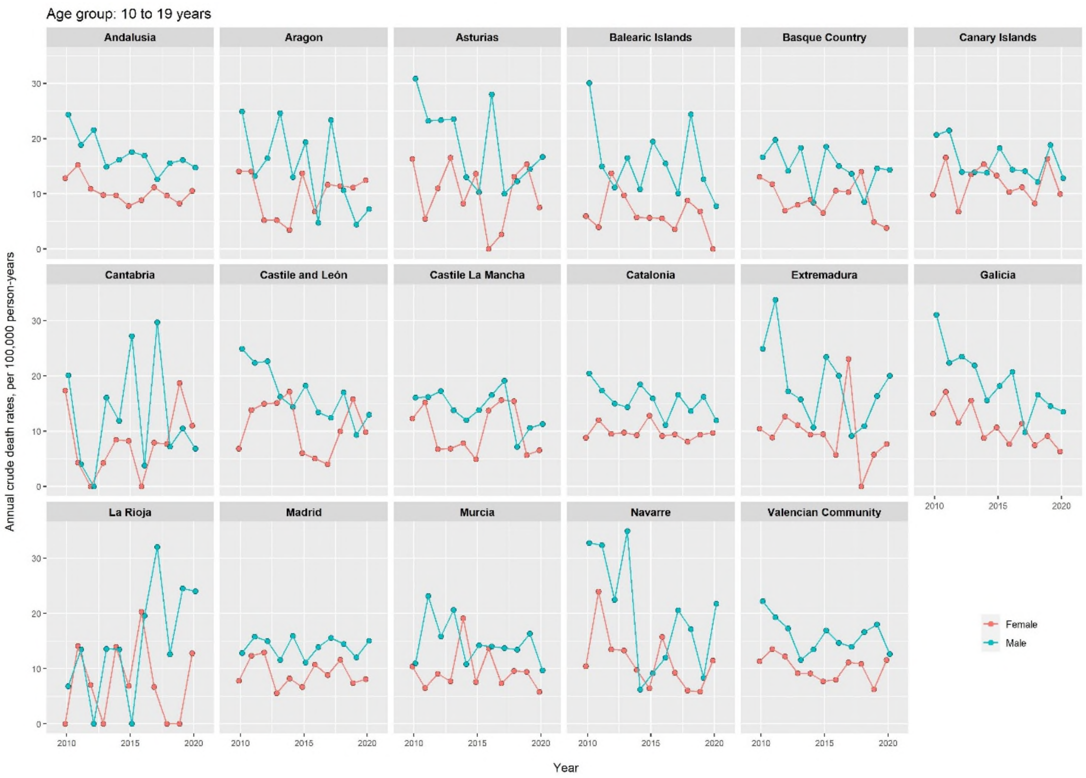

S1 (C): 20-29 years

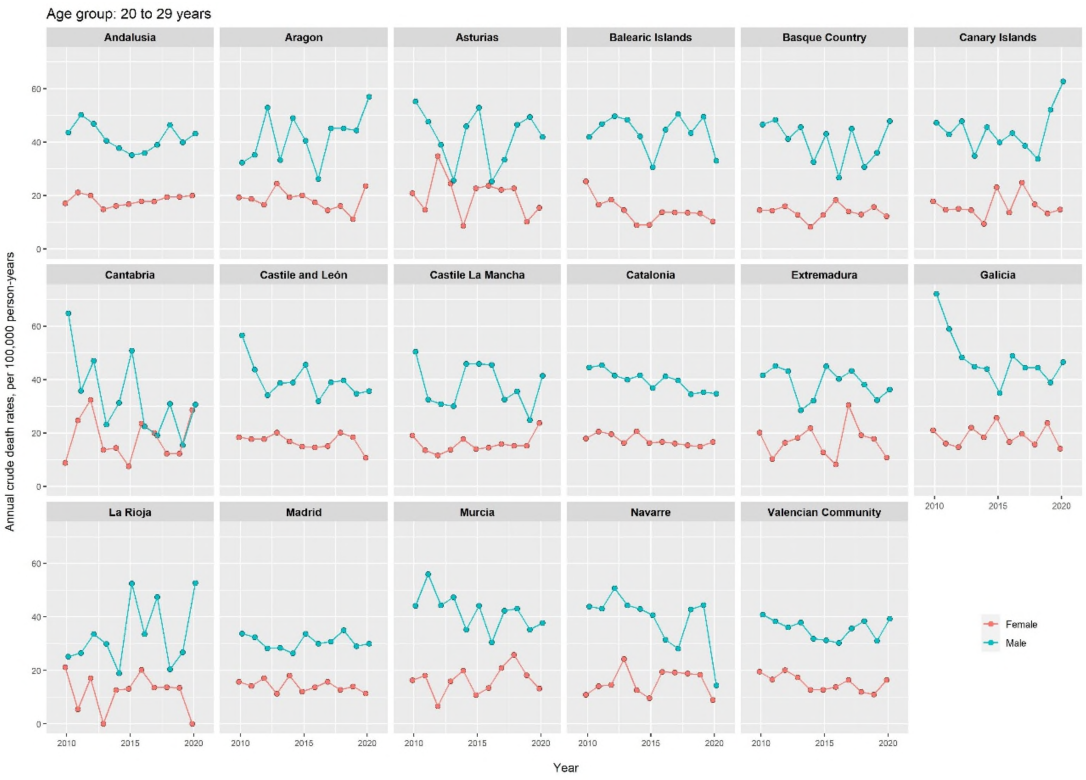

### S1 (D): 30-39 years

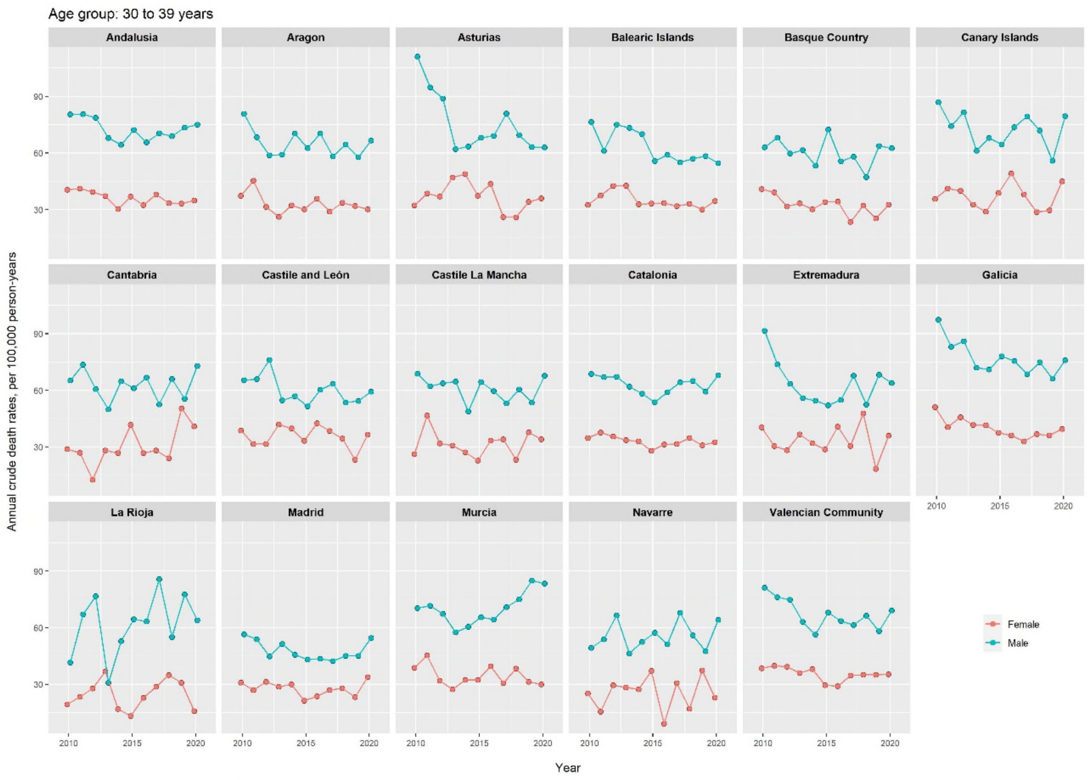

S1 (E): 40-49 years

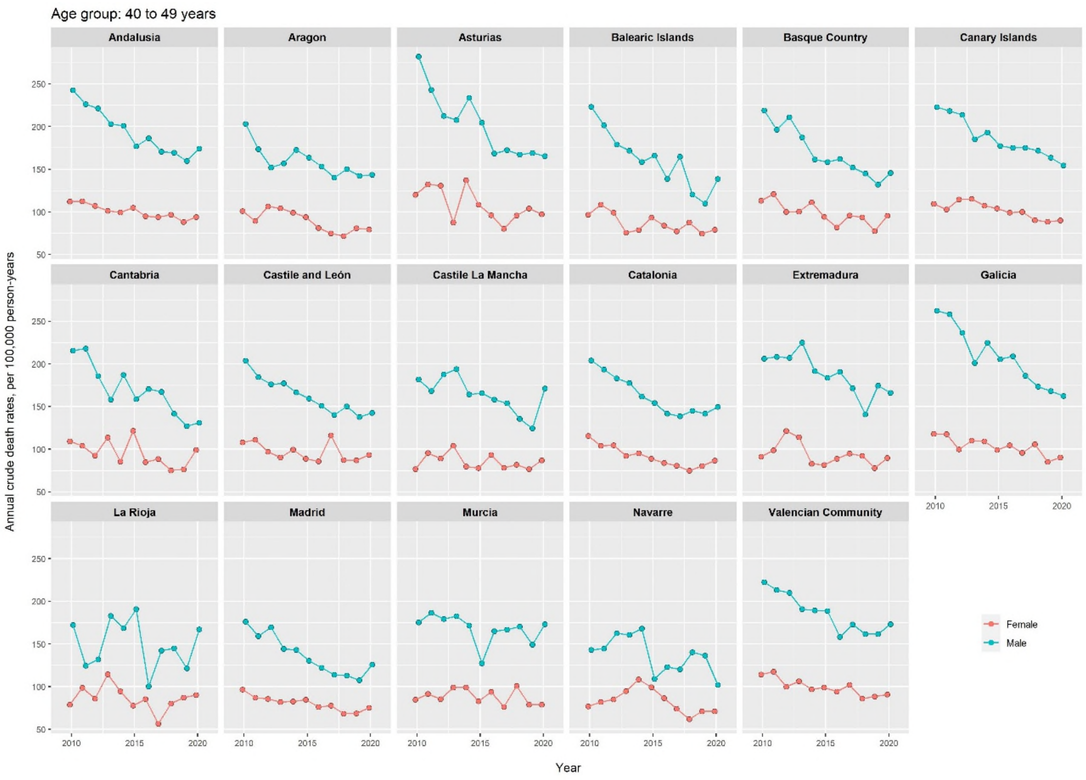

S1 (F): 50-59 years

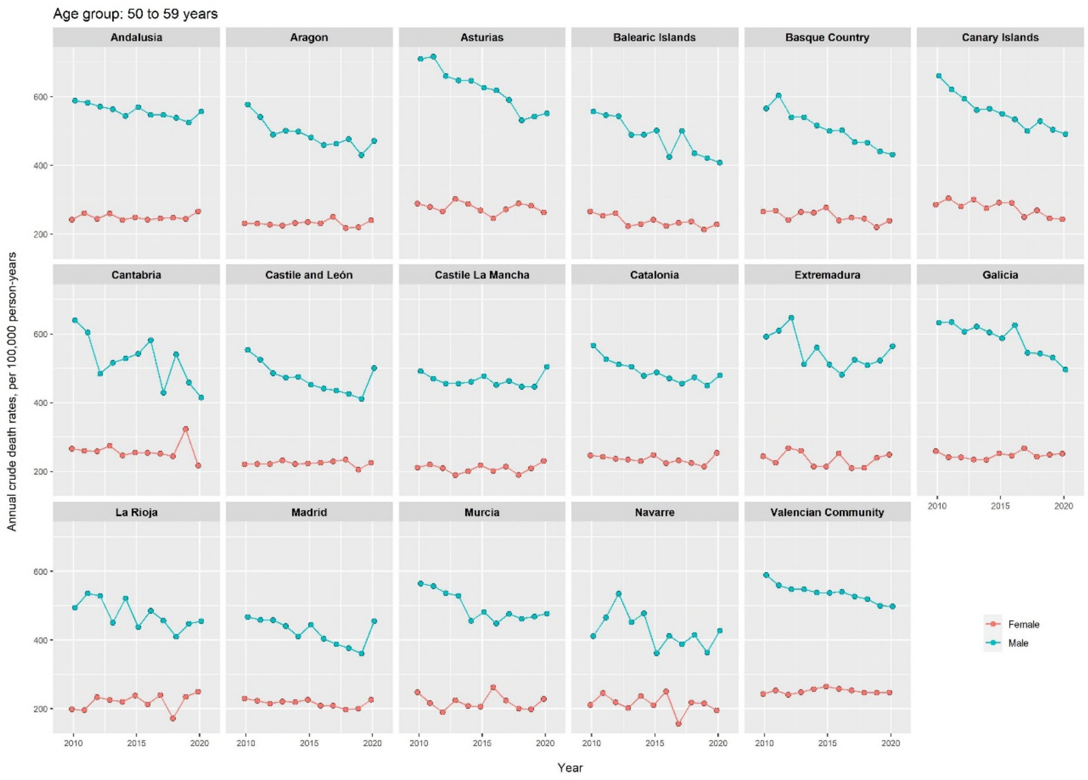

S1 (G): 60-69 years

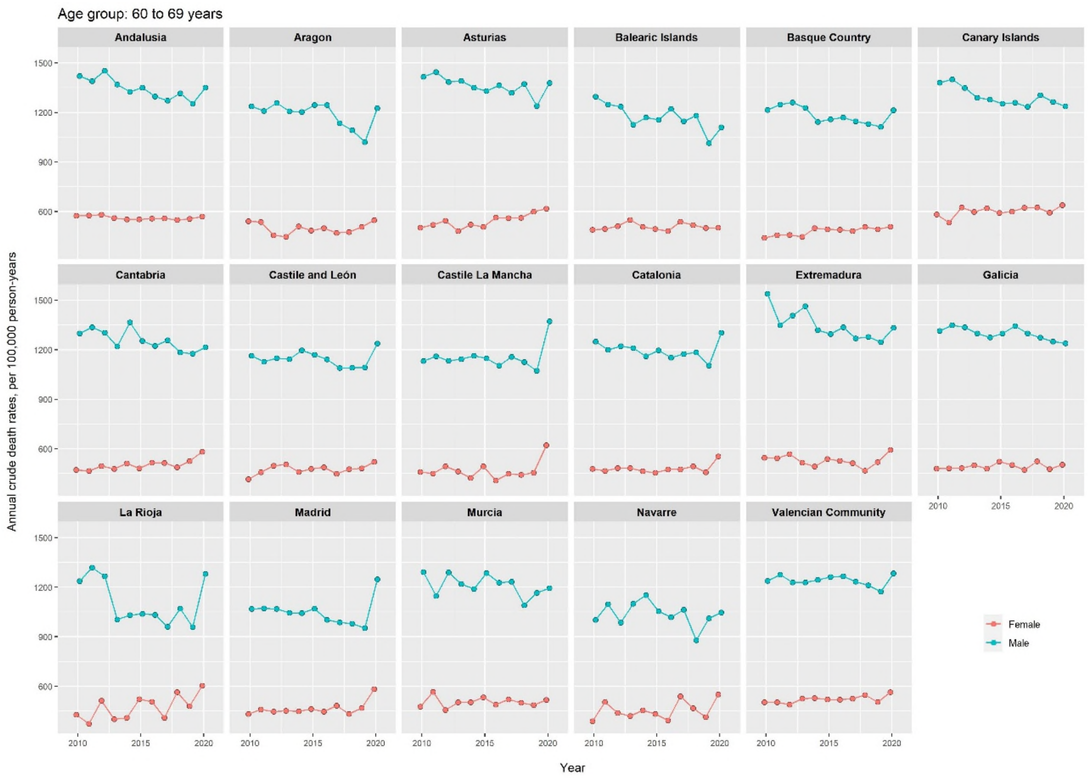

S1 (H): Sex difference (men minus women) in age-standardised annual mortality rate (per 100,000)

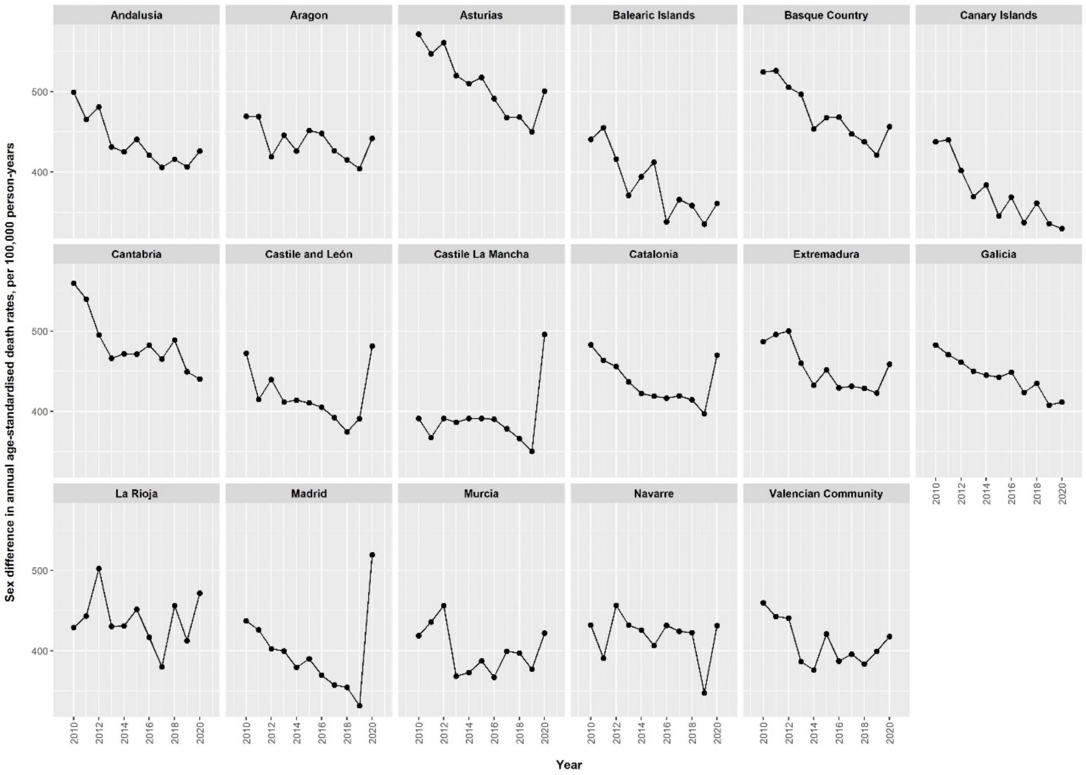

Supplementary Figure S2: Crude and age-standardised excess death rates in Spanish regions during 2020-2021, by sex

S2 (A): Crude and age-standardised excess death rates

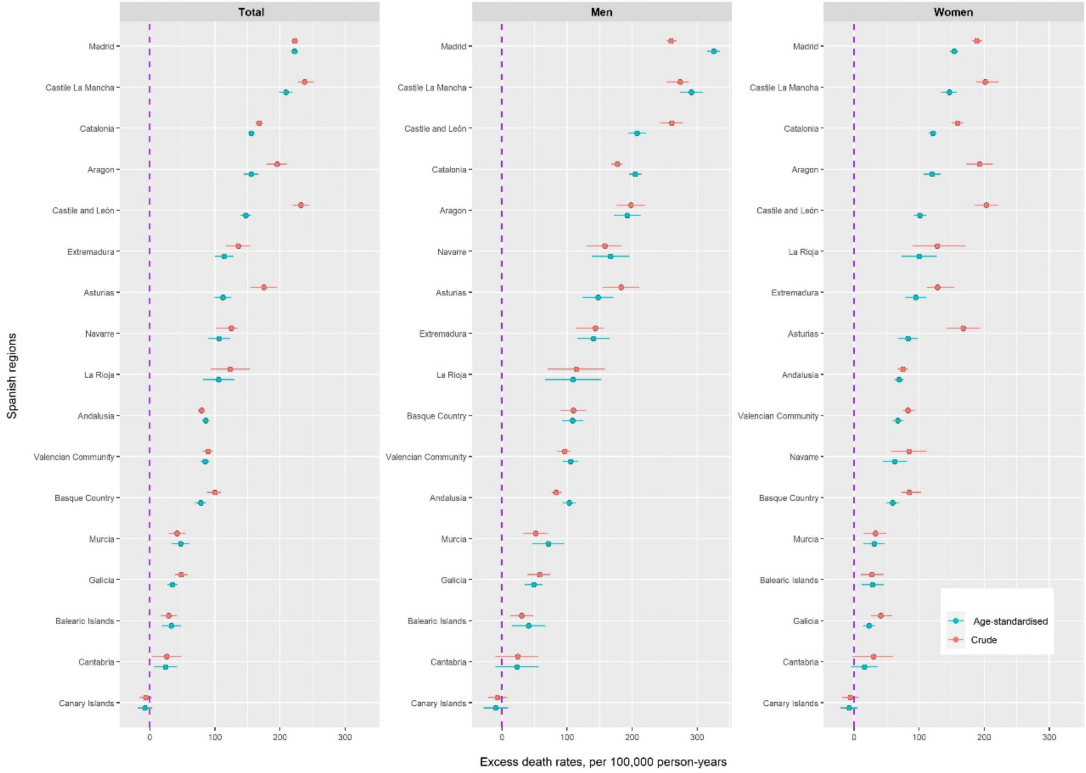

S2 (B): sex differences in crude and age-standardised excess death rates

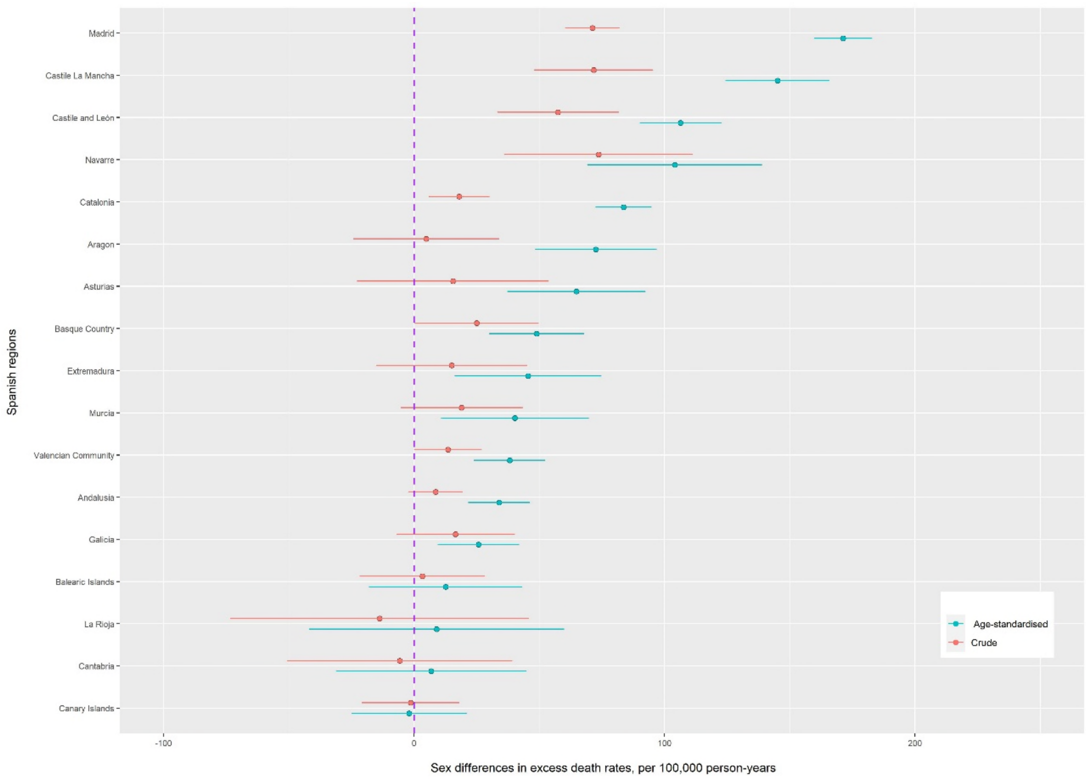

-Standandardised using 2013 European standard population. Confidence intervals were estimated assuming a Poisson distribution of the mortality rates.

Supplementary Figure S3: Years of life lost in Spanish regions, 2010-2020

S3 (A): Trend of years of life lost, by sex

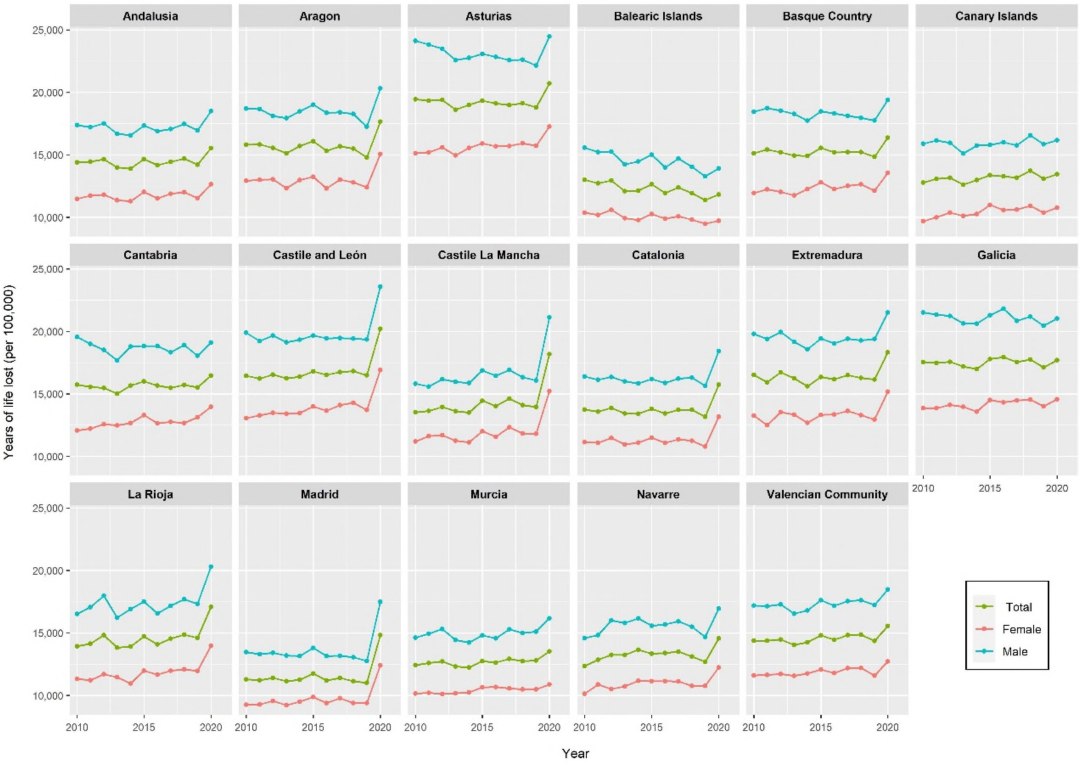

S3 (B): Trend of years of life lost by age groups in men

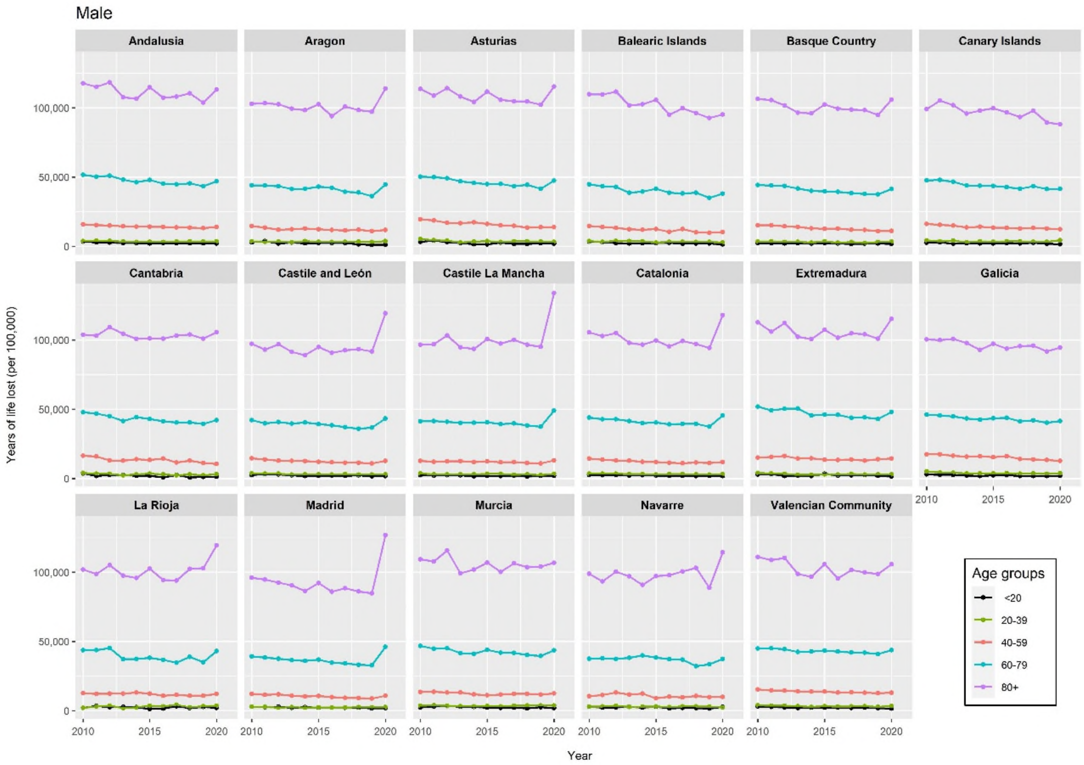

S3 (C): Trend of years of life lost by age groups in women

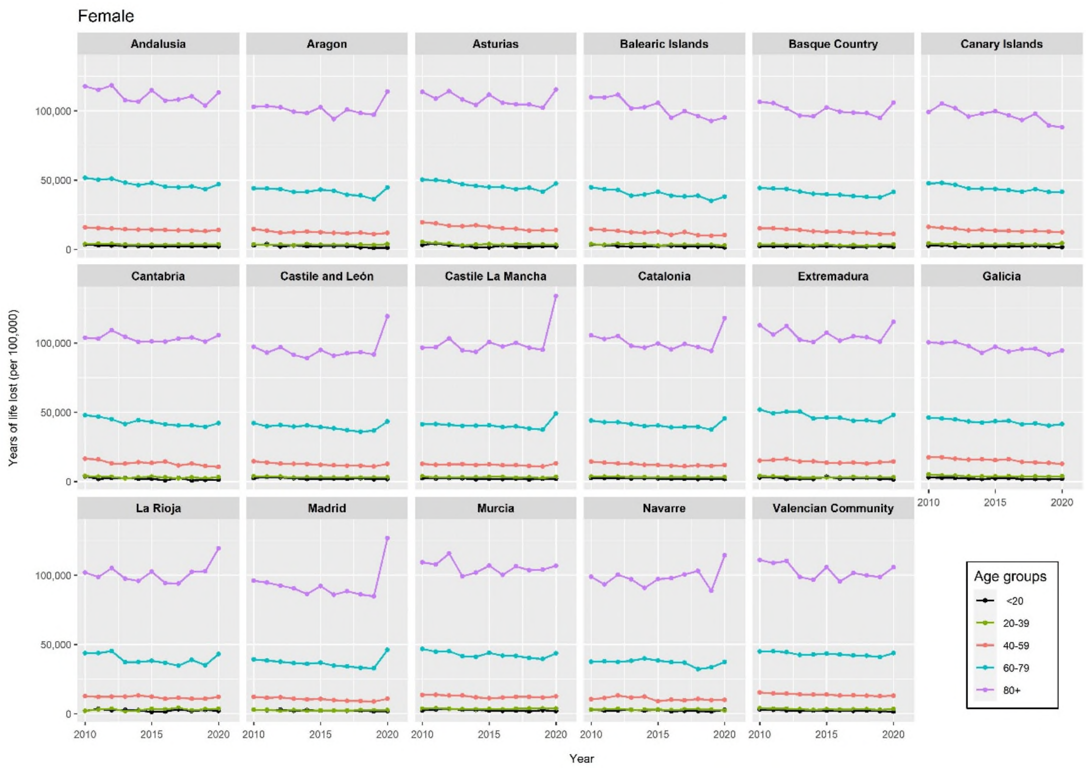

S3 (D): Changes in years of life lost in 2020, by sex

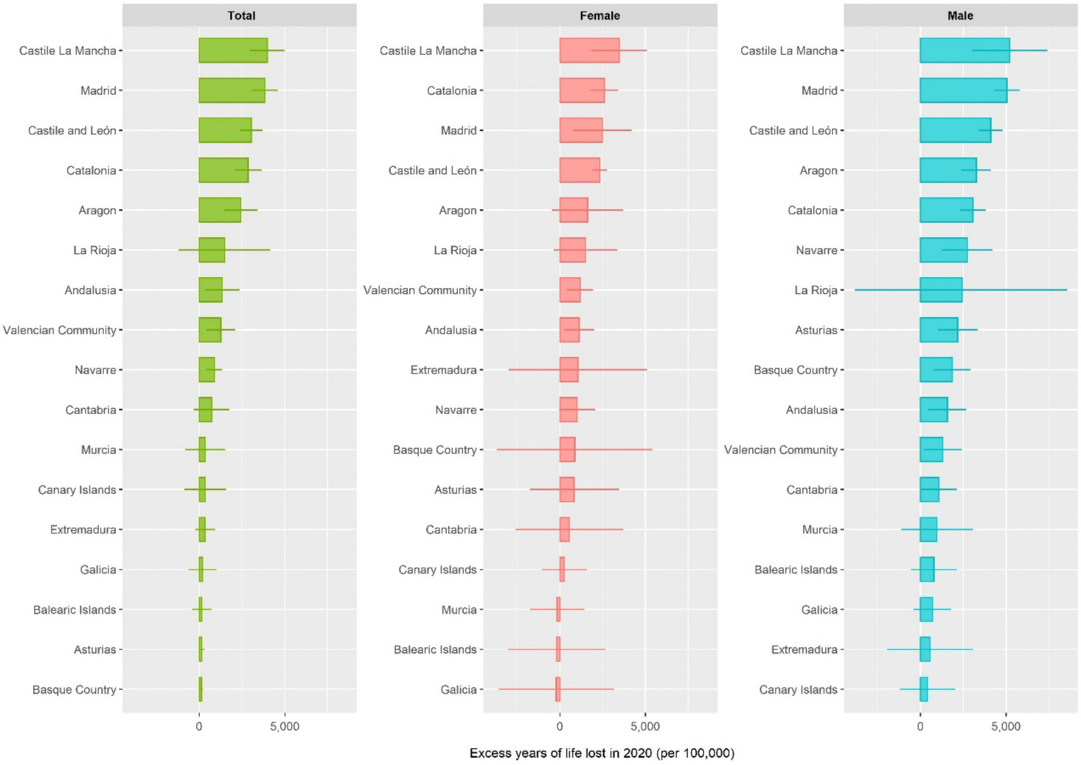

S3 (E): Changes in years of life lost in 2020, by age and sex

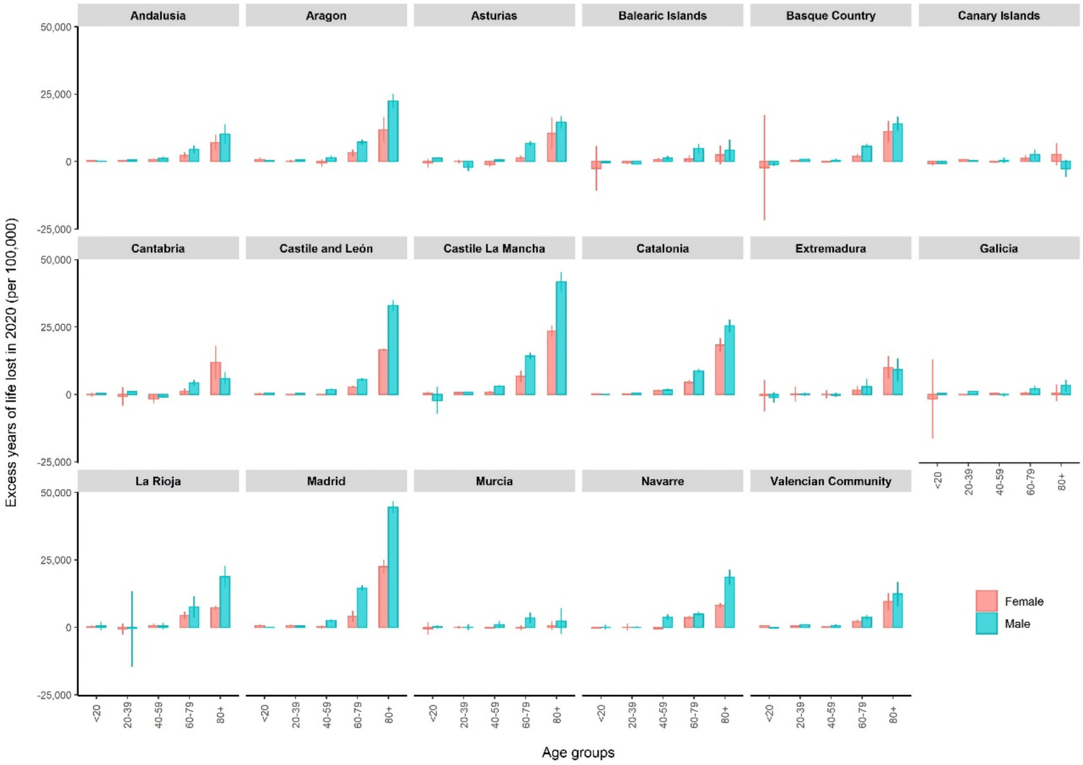
